# Genomic Determinants of Biological Age Estimated By Deep Learning Applied to Retinal Images

**DOI:** 10.1101/2024.09.18.24313871

**Authors:** Yu Huang, Mohammad Ghouse Syed, Ruiye Chen, Cong Li, Xianwen Shang, Wei Wang, Xueli Zhang, Xiayin Zhang, Shulin Tang, Jing Liu, Shunming Liu, Sundar Srinivasan, Yijun Hu, Muthu Rama Krishnan Mookiah, Huan Wang, Emanuele Trucco, Honghua Yu, Colin Palmer, Zhuoting Zhu, Alexander S F Doney, Mingguang He

## Abstract

With the development of deep learning (DL) techniques, there has been a successful application of this approach to determine biological age from latent information contained in retinal images. Retinal age gap (RAG) defined as the difference between chronological age and predicted retinal age has been established previously to predict the age-related disease. In this study, we performed discovery genome-wide association analysis (GWAS) on the RAG using the 31271 UK Biobank participants and replicated our findings in 8034 GoDARTs participants. The genetic correlation between RAGs predicted from the two cohorts was 0.67 (P=0.021). After meta-analysis, we found 13 RAG loci which might be related to retinal vessel density and other aging processes. The SNP-wide heritability (h2) of RAG was 0.15. Meanwhile, by performing Mendelian Randomization analysis, we found that glycated hemoglobin, inflammation hemocytes and anemia might be associated with accelerated retinal aging. Our study explored the biological implications and molecular level mechanism of RAG, which might enable causal inference of the aging process as well as provide potential pharmaceutical intervention targets for further treatment.

## Introduction

Individuals vary in their susceptibility to degenerative changes associated with age^1^. Biological age (BA) or functional age may be a more important determinant of age-related health problems than chronological age (CA). Conventionally, BA can be defined by a range of hallmarks such as telomere length, epigenetic alterations, DNA methylation or mitochondrial function^2,3^. Estimated BA derived from such biological markers outperformed CA in predicting frailty and mortality^4^ suggesting its superiority in predicting general age-related health outcomes^5^.

Recently, with the development of deep learning (DL) techniques, there has been growing interest in the use of this approach to determine BA from latent information contained in medical images. For example, BA derived from brain magnetic resonance images (MRI)^6,7^, from liver and pancreas MRI^8^ and chest radiographs^9^. These BAs have shown good performance in predicting overall mortality. However, according to the American Federation of Aging Research (AFAR)^10^, a biomarker of aging should ideally be more efficient and accurate than CA in predicting longevity meanwhile remain applicable to general diseases and be widely practicable and cost-effective. Thus, effective, non-specific, non-invasive and economic aging biomarkers were still needed.

The retina comprises a highly vascularized neurological tissue and is widely regarded as a window indicating global tissue health. For example, retinal vascular features are a strong predictor for all-cause mortality and other cardiovascular or kidney diseases^11,11^; while retinal neuron tissue can also be a good marker for systemic neuron-degenerative diseases^15,16^. Importantly, highly detailed images can be conveniently acquired with relatively low-cost equipment and minimal training. This makes images of the retina highly attractive as a substrate for deep learning to investigate ageing.

Currently, several studies have reported a DL approach to predict age using retinal images. Preliminary findings suggested that the predicted retinal age gap (RAG, the difference between chronological age and predicted retinal age) was strongly predictive of mortality and a range of other conditions **^17,17^**. However, only one study explored its biological (genetic) causal determinants**^20^** but without further validation. The biological implications of RAG should be fully explored in order to enable causal inference of the aging process. Furthermore, a molecular level mechanistic understanding of RAG might ultimately make it possible to apply pharmaceutical interventions to achieve its beneficial effects.

It is likely that the rate of tissue ageing is a consequence of both endogenous genetic factors and environmental exposures. To further improve the understanding of the RAG, we conducted a genome-wide association study (GWAS) on RAGs obtained from two distinct populations, UK Biobank and GoDARTS with different DL models. Firstly we tested the genetic concordance of RAGs derived from different DL algorithms; Secondly, we explored the biological relevance of this trait; Third we determined biological causal factors in a Mendelian Randomization (MR) framework.

## Results

### Stage 1: Discovery of RAG risk loci in the UK Biobank

In stage 1, we performed GWAS in the UK Biobank data set. A total number of 31271 European participants were involved and 6133594 SNPs were tested. After Bonferroni correction and SNP clumping, 16 sentinel SNPs reached genome-wide significant level. The strongest signal was found for SNP rs60149603 (P=1.4e-95) located on chromosome 2, and 7 independent further SNPs were also identified from the same chromosome. The second strongest signal was rs8001273 (5.7e-22) located on chromosome 13 within the gene LOC105370101. Other SNPs are provided in Supplementary Table 1. The overall heritability estimated by LDSC was 13.8% and there was no evidence of inflation due to population stratification (LDSC intercept 1.01, se = 0.01). GWAS for the non-transformed RAG and the logistic model demonstrated similar findings (Supplementary Figure 1 a, b, c).

### Stage 2: Replication of the genetic effect in the GoDARTS cohort

Stage 2 analysis was performed in GoDARTS. After meta-analysis of different phases, 8034 participants with 8625586 SNPs were analyzed. Among the 16 sentinel loci in the UK Biobank study, 2 SNPs were not found in the GoDARTS study and 7 SNPs were replicated in the GoDARTS stage 2 analysis with P < 0.01. The effect of these SNPs was in the same direction (Supplementary Table 1). There was a relatively high concordance between the two populations for the significant loci. The Pearson correlation coefficient of the effect sizes is (r) = 0.82 (Figure 2). For the overall genetic effect, we confirmed a high genetic correlation between RAG in UK Biobank and RAG in GoDARTS (rg=0.67, P=0.021). As a sensitivity analysis, we also performed a GWAS of RAG for the UK Biobank T2D population (3279 participants) and we had similar findings (Supplementary Table 1).

**Figure 1.**
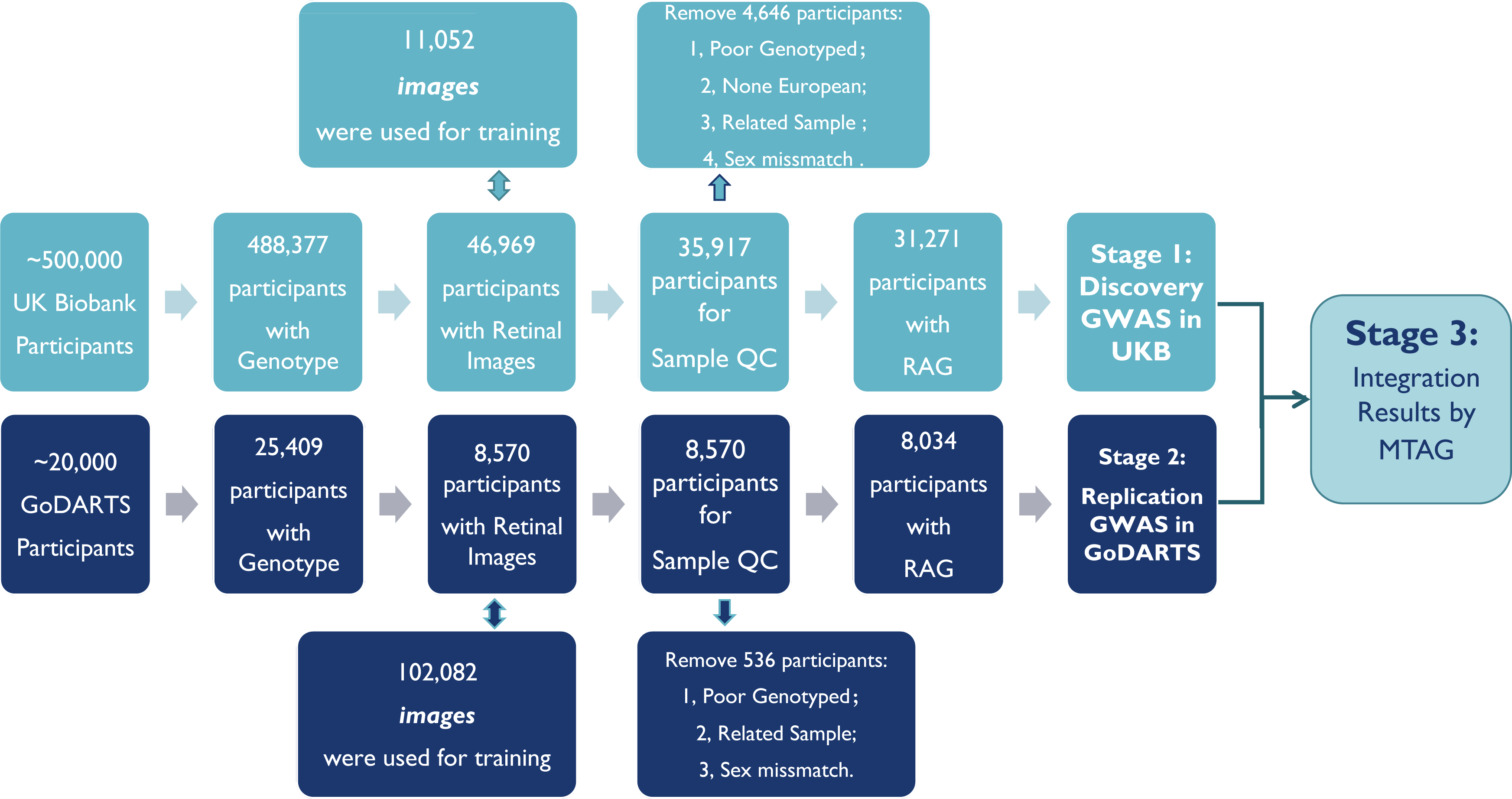
Flowchart of the study design. This figure summarizes the three stages of this study, as well as the data resources and main quality control process for each stage.

**Figure 2.**
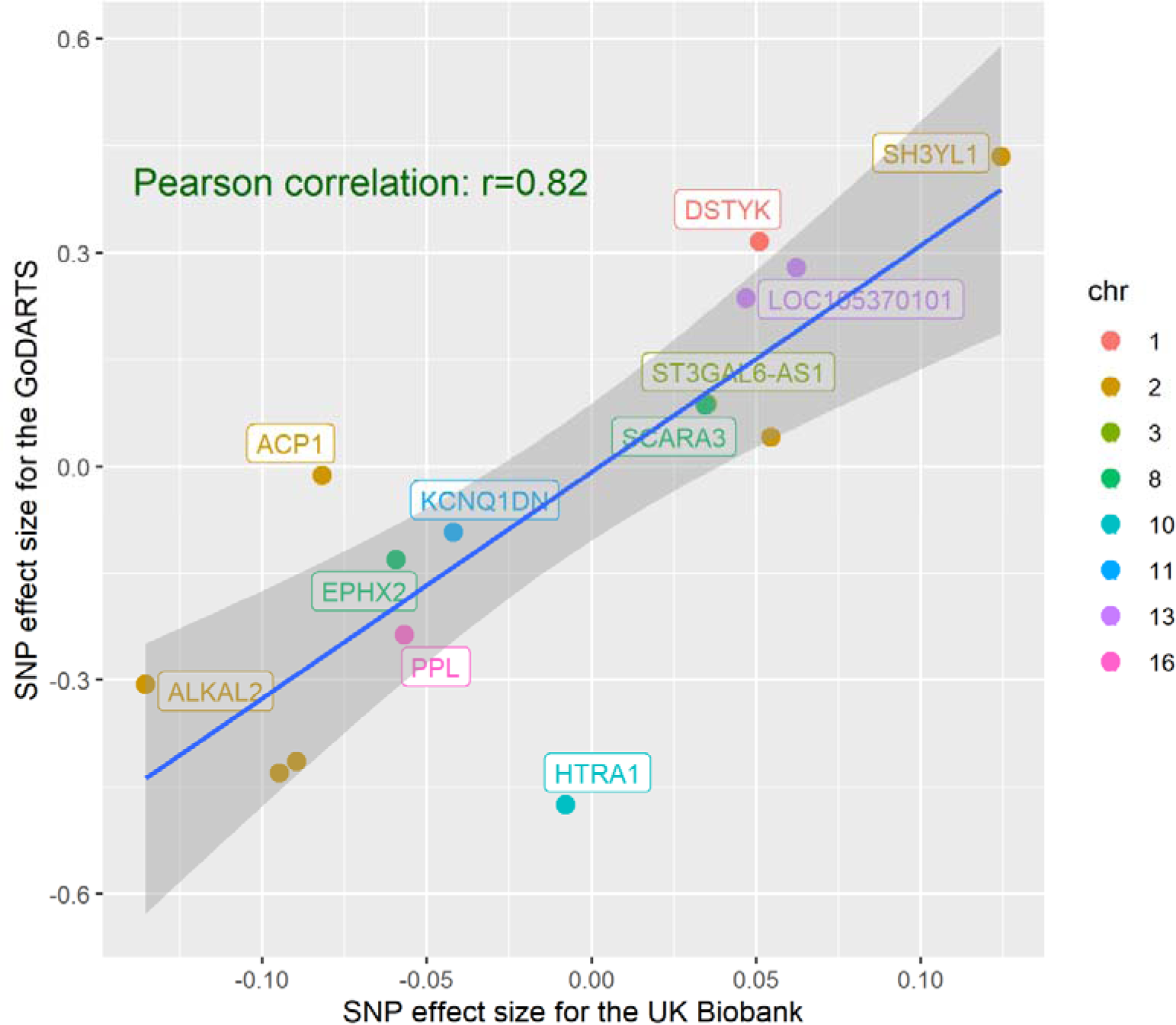
Pearson correlation of the effect sizes of the top SNPs derived from two different GWASs of RAG. The x-axes show RAG SNP effect estimates for the stage 1 GWAS using the UK Biobank population. The y-axes show the effect estimates for the same SNPs obtained from the stage 2 GWAS using the GoDARTS population. Eeach dot represent a SNP and the colour represent SNPs from different chromosomes.

Apart from replicating the findings from stage 1, GWAS of RAG in the GoDARTS identified additional SNPs: one SNP rs932275 (p=3.04E-10) on chromosome 10 located in the intron region of HTRA1 – a gene previously associated with age-related macular degeneration (AMD) and second SNP, rs13274427 (P = 1.35E-08) on chromosome 8 located in KBTBD11/KBTBD11-OT1 reached a genome-wide significant threshold. However, neither of these SNPs was nominally significant in the UK Biobank (Supplementary Figure 2, supplementary table 1). The overall heritability was slightly lower than the UK Biobank cohort with an estimated h2 of 10.1% while the inflation remained low (LDSC intercept 1.01, se = 0.007).

### Stage 3: Discovery of novel RAG loci via Multi-trait genome-wide association analysis (MTAG)

The MTAG yield 13 statistically independent sentinel SNPs (Table 1, Figure 3) from 8 leading genomic regions (Supplementary figure 3). This included all previously UK Biobank regions as well as 1 novel region from chromosome 1 which encodes *DSTYK* (Supplementary table 1). Joint and conditional analysis by cojo-GCTA didn’t identify any further independent signals. As expected, the overall heritability was increased (h2=15%), and there was no sign of inflation (intercept = 1.00, se=0.01).

**Figure 3.**
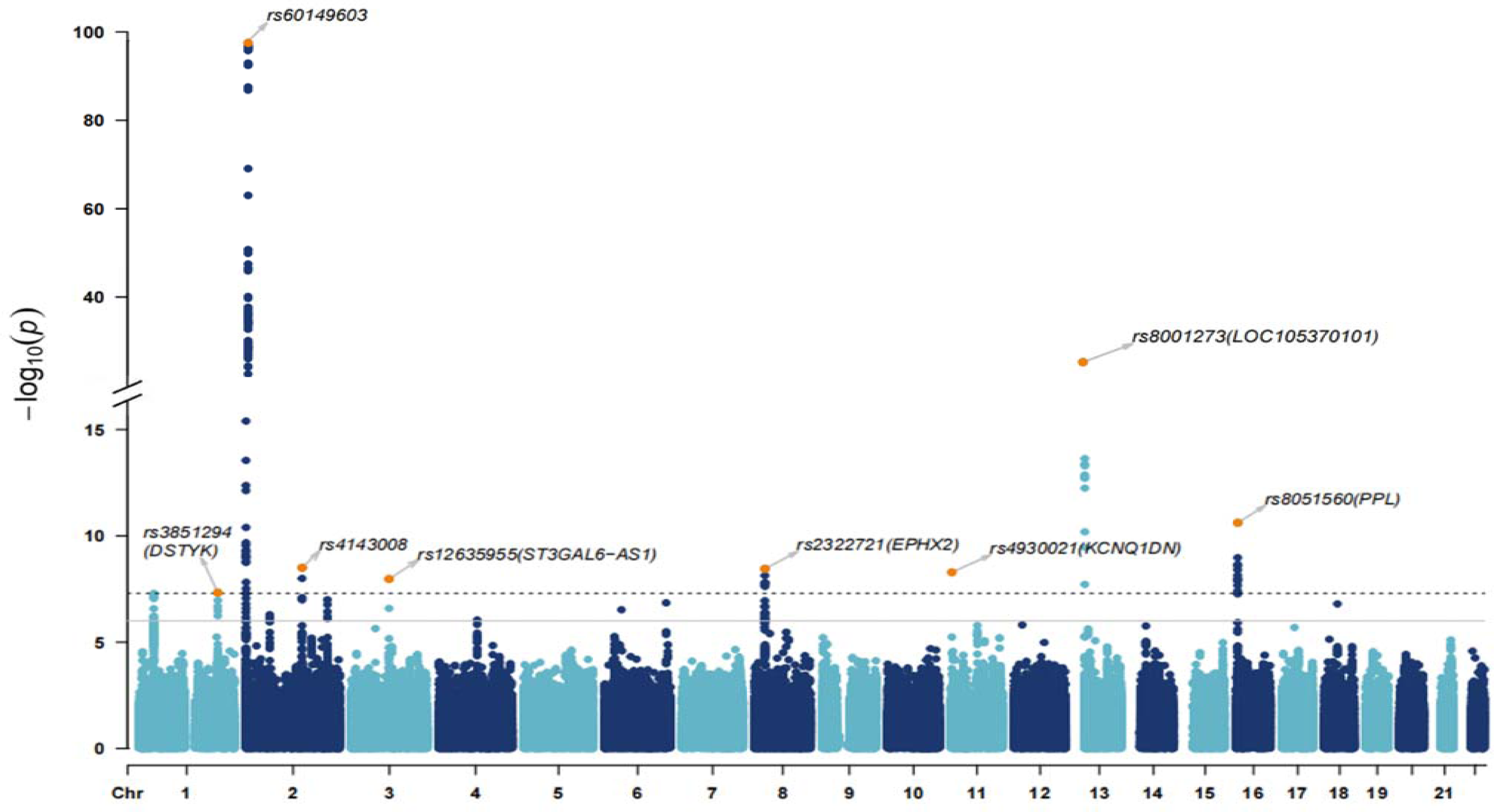
Manhattan plot displaying RAG P values from the multi-trait GWAS (MTAG) analysis. Each dot represents a SNP, the x-axis shows the chromosomes where each SNP is located, and the y-axis shows −log10 P-value of the association of each SNP with RAG in the stage 3 MTAG analysis. The dash horizontal line shows the genome-wide significant threshold (P-value = 5e-8) and the solid gray line shows the suggestive significant threshold (P-value = 1e-5). The nearest gene to the most significant SNP in each locus has been labeled.

**Table 1:**
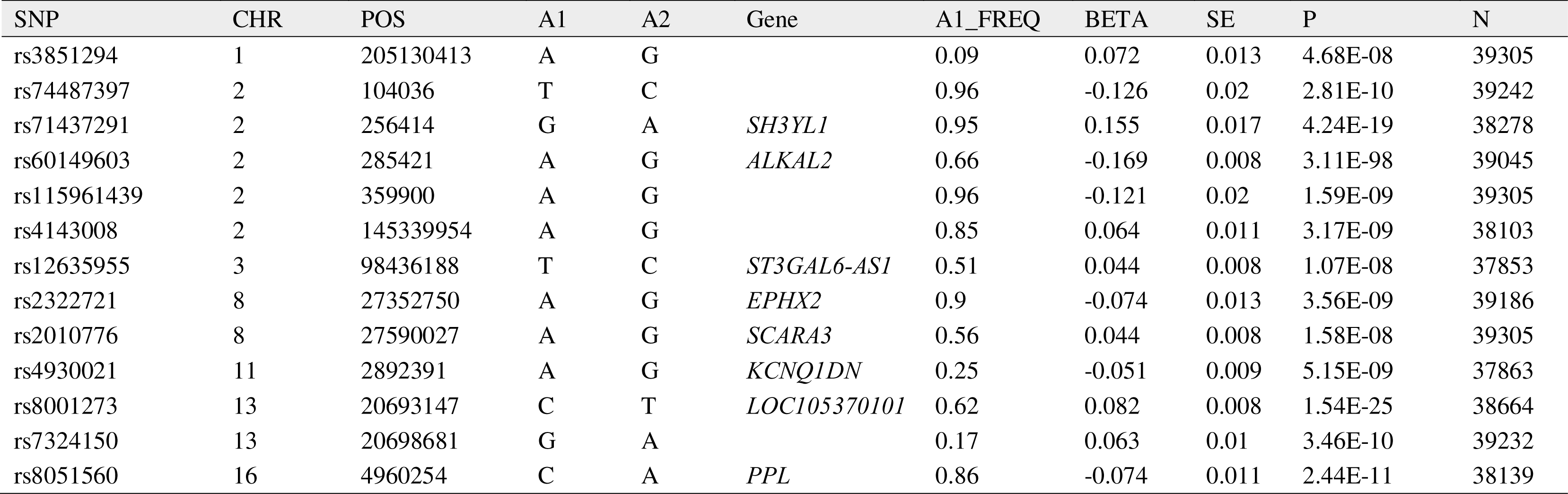
RAG loci via stage 3 MTAG analysis.

| SNP | CHR | POS | A1 | A2 | Gene | A1_FREQ | BETA | SE | P | N |
| --- | --- | --- | --- | --- | --- | --- | --- | --- | --- | --- |
| rs3851294 | 1 | 205130413 | A | G |  | 0.09 | 0.072 | 0.013 | 4.68E-08 | 39305 |
| rs74487397 | 2 | 104036 | T | C |  | 0.96 | -0.126 | 0.02 | 2.81E-10 | 39242 |
| rs71437291 | 2 | 256414 | G | A | *SH3YL1* | 0.95 | 0.155 | 0.017 | 4.24E-19 | 38278 |
| rs60149603 | 2 | 285421 | A | G | *ALKAL2* | 0.66 | -0.169 | 0.008 | 3.11E-98 | 39045 |
| rs115961439 | 2 | 359900 | A | G |  | 0.96 | -0.121 | 0.02 | 1.59E-09 | 39305 |
| rs4143008 | 2 | 145339954 | A | G |  | 0.85 | 0.064 | 0.011 | 3.17E-09 | 38103 |
| rs12635955 | 3 | 98436188 | T | C | *ST3GAL6-AS1* | 0.51 | 0.044 | 0.008 | 1.07E-08 | 37853 |
| rs2322721 | 8 | 27352750 | A | G | *EPHX2* | 0.9 | -0.074 | 0.013 | 3.56E-09 | 39186 |
| rs2010776 | 8 | 27590027 | A | G | *SCARA3* | 0.56 | 0.044 | 0.008 | 1.58E-08 | 39305 |
| rs4930021 | 11 | 2892391 | A | G | *KCNQ1DN* | 0.25 | -0.051 | 0.009 | 5.15E-09 | 37863 |
| rs8001273 | 13 | 20693147 | C | T | *LOC105370101* | 0.62 | 0.082 | 0.008 | 1.54E-25 | 38664 |
| rs7324150 | 13 | 20698681 | G | A |  | 0.17 | 0.063 | 0.01 | 3.46E-10 | 39232 |
| rs8051560 | 16 | 4960254 | C | A | *PPL* | 0.86 | -0.074 | 0.011 | 2.44E-11 | 38139 |

### Functional annotation of the sentinel 13 SNPs identified by MTAG

For each of the sentinel loci, SNPs that were in LD (r2 ≥ 0.8) were annotated, and all SNPs were mapped to the nearest gene within 10kb of the SNP. The majority of the SNPs were intergenic and 28 gene regions were mapped. Two of the sentinel SNPs -rs3851294 on chromosome 1 and rs4143008 on chromosome 8-may have deleterious effects (CADD scores >12.37), 79% of these SNPs are expression quantitative trait loci (eQTLs) and 2016 chromatin interactions with the 8 leading regions were observed. (Supplementary results 1, supplementary table 2-3 and supplementary figure 4-5).

### In silico look up for pleiotropic effects

By looking up the SNPs in PhenoScanner and GWAS catalog, we evaluated the cross-trait and disease associations of the sentinel SNPs identified from MTAG. By selecting traits at a genome-wide significant level (5e-08), the search of published GWAS showed that 7 of our 13 loci (using sentinel SNPs or proxies estimated in the European population; r2 ≥ 0.8) were also associated with a wide range of traits and diseases. Among them, anthropomorphic traits such as BMI, and body height measures were most commonly shared with RAG SNPs, highlighting a common link between body growth and RAG. Haematological measurements such as mean corpuscular hemoglobin concentration, white blood cell and Basophil count were also associated with different RAG SNPs, indicating the potential importance of cellular blood constituents in retinal aging. Ophthalmic measurements such as spherical equivalent were also tagged by proxy SNPs. In particular, rs3851294 in *DSTYK* and rs60149603 in *ALKAL2* demonstrated variety of pleiotropic effects. These findings suggest a complex array of biological processes involved in retinal aging (Figure 4, Supplementary Table 4).

**Figure 4.**
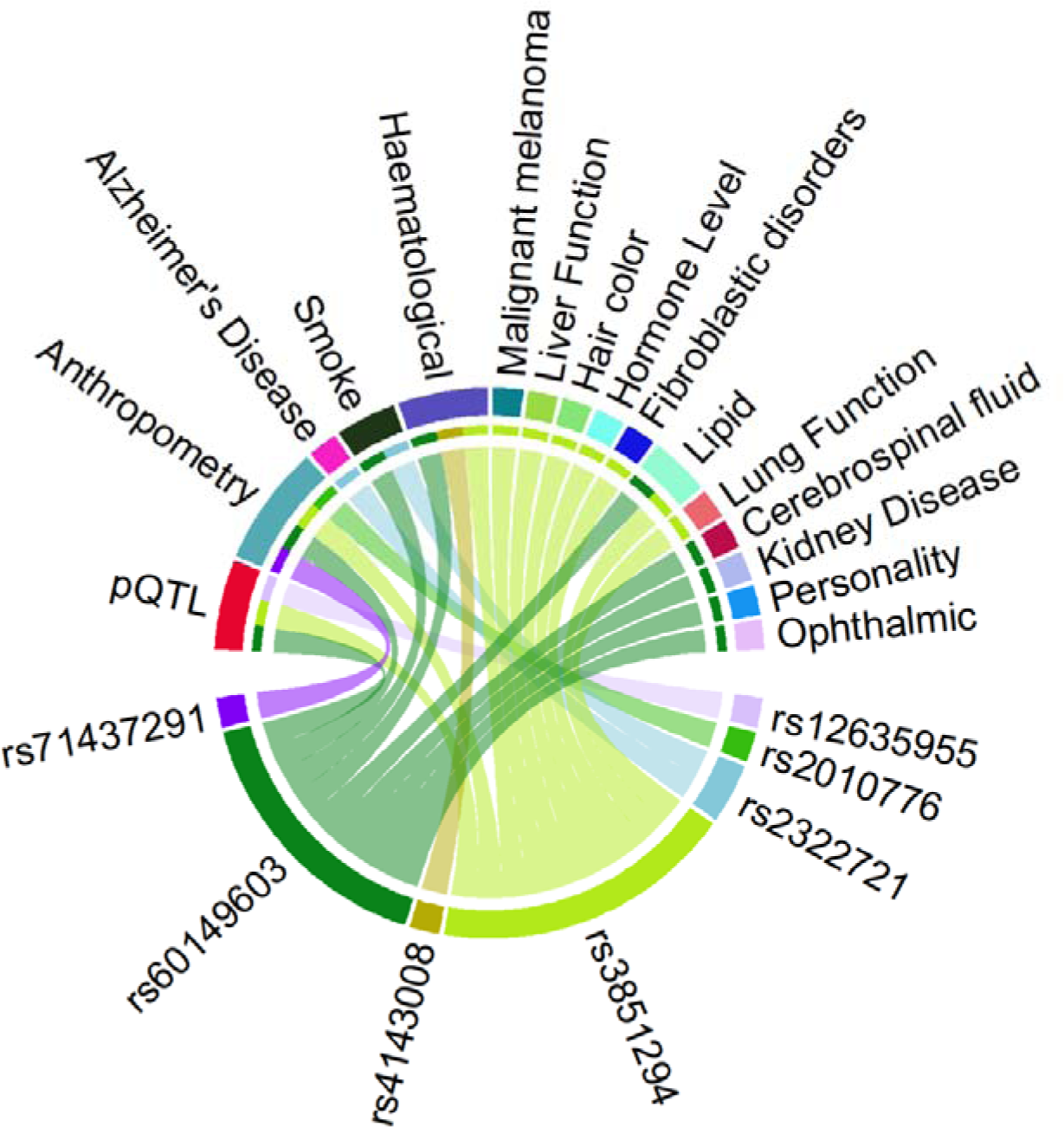
Association of RAG loci with different traits. Plot shows results from associations with other traits which were extracted from the PhenoScanner / GWAS catalog databases for the 14 novel sentinel SNPs including proxies in Linkage Disequilibrium (r2 ≥ 0.8) with genome-wide significant associations.

We further explore the association of sentinel SNPs with lifestyle traits by looking up the PheWAS in the UK Biobank via the Gene ATLAS database (N=408,455). At a moderate level (P < 0.05), we found genetic associations of RAG variants with different lifestyle traits. However, the overall associations demonstrated heterogeneous effects with life style. More details are in supplementary results 2 supplementary table 5 and supplementary figure 6.

### Enrichment analysis

We performed gene-based and pathway-based tests using MAGMA v1.07b in stage 1 and 2 results separately, and lastly in the MTAG result (Supplementary table 6 and supplementary figure 7 -c demonstrated the prioritised genes of each stage). The genome-wide significance level was defined at P =2.607e-6 (0.05/19182). Two genes *SH3YL1* and *ACP1* located on chromosome 2 were prioritized in each GWAS stage 1, 2 and 3. *SH3YL1* is a newly identified retinal vessel density gene^12^, indicating retinal vascular features might also be a feature of RAG. We found two genes -ARMS2 and PLEKHA1-located on chromosome 10 were enriched in GoDARTS data and had P values of 7.07E-04 and 0.03 in MTAG. Though not reaching genome-wide significance these genes were AMD genes, suggesting that macular degeneration might be a further feature of RAG (Supplementary figure 7 b). Overall, 9 genes were prioritized for MTAG results, among them, *TMCC2* from chromosome 1 and *CCDC25* from chromosome 2 were newly identified genes (Supplementary table 6, supplementary figure 7).

None of the enriched pathways was significant after adjustment (FDR < 0.05). Enrichment of the MTAG result suggests a reactome pathway - ‘melanin biosynthesis’ - might be involved in the retinal aging process (P=1.17E-05). The top pathways were shown in supplementary table 7. Both MAGMA and hypergeometric mean pathway analysis (GENE2FUNC implemented in FUMA) didn’t identify any enrichment of the RAG gene in any tissue. The overall expression pattern of the mapped genes in different tissues and cells was shown in supplementary figure 8 and 9.

### Different biological ages share common genes

For further validation, we compared the genes identified in our study (from stages 1, 2 and 3) with genes of different biological ages identified by Goallec et.al.^20^. In Goallec et.al.’s study, RAG was calculated based on eye front segment image, fundus image or OCTA. We note that, among the 30 unique genes reported in our study (either from direct mapping or from enrichment analysis), 13 of them were overlapping with the RAG genes reported by Goallec et.al. Especially, 10 genes (*ACP1, SH3YL1, FAM110C, OCA2, CCDC25, PPL, MACF1, ST3GAL6-AS1, UBN1, EPHX2*) were exactly matching with RAG genes derived from retinal fundus images (Figure 5 a). Moreover, 8 of our RAG genes were shared by other biological ages: HTRA1 was identified as genes for both ‘abdominal age’ and ‘lungs age’ while *ARMS2 and PLEKHA1* were genes for ‘abdominal age’ and *PPL*, *ST3GAL6-AS1, ROGDI* and *MACF1* were ‘lungs age’ genes; *SH3YL1* and *ACP1* were identified as genes for ‘biochemistry age’ and *TMCC2 and DSTYK* were also a gene for ‘blood cells age’ (Figure 5 b).

**Figure 5.**
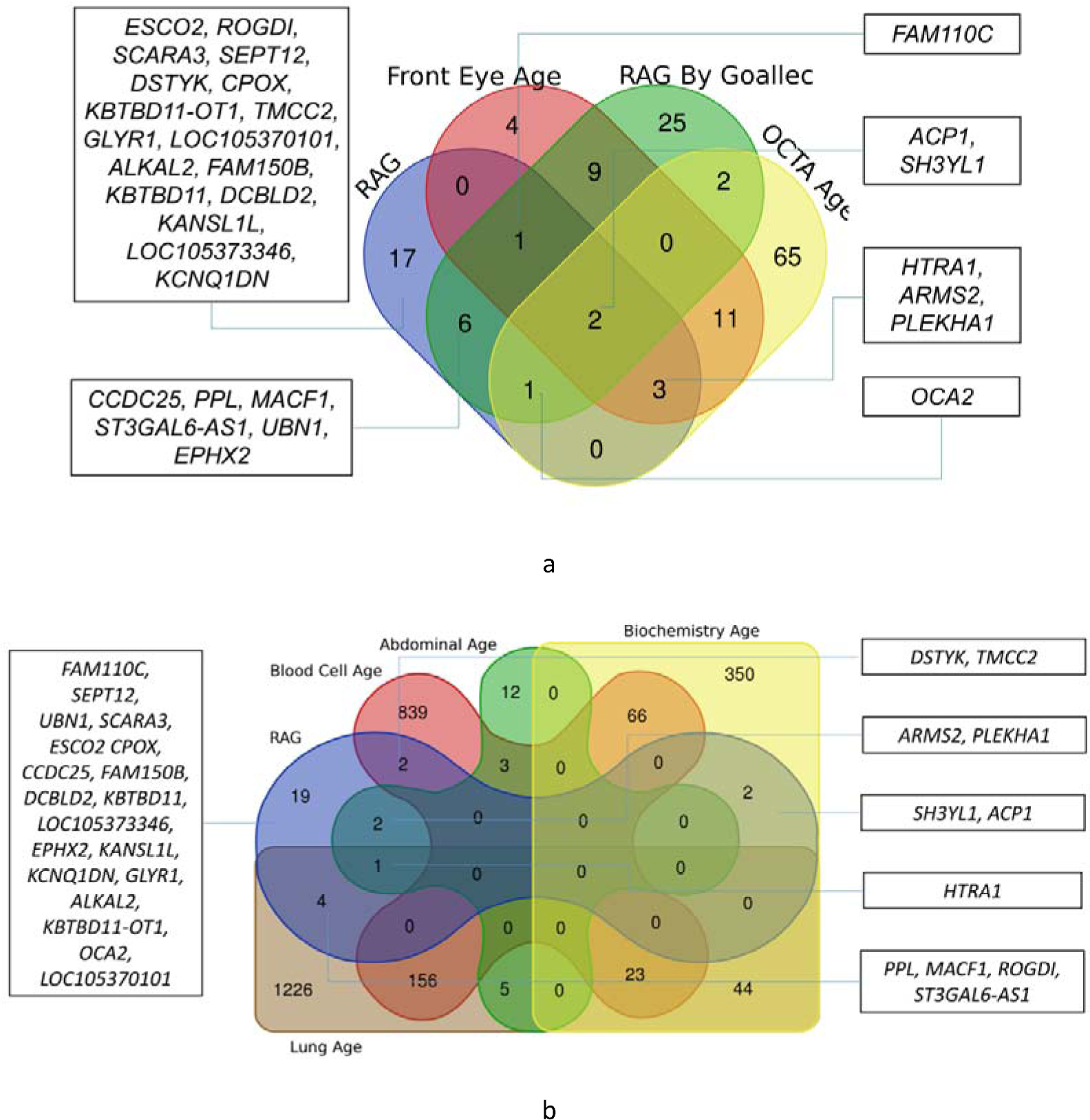
Venn-diagram of overlapped genes of different biological ages. Genes for different BAs were reported by Goallec et.al^20^, data were derived from https://www.multidimensionality-of-aging.net/. A) Common genes of RAG between our study and Goallec’s findings. B) Common genes between RAG and other biological age.

### Genetic correlation analysis

We further applied linkage disequilibrium score regression analysis to test the genetic correlation between RAG and multiple ocular and non-ocular traits. However, none of the results passed the Bonferroni multiple testing. Details were listed in supplementary table 8.

### Mendelian Randomization analysis: HbA1c and blood lipid, refractive status, inflammation related hemocytes might cause retinal aging

We performed a series of two-sample Mendelian randomization studies to assess the putative causal relationships between systemic phenotypes on RAG. To select instrumental variables (IV), for each trait, we removed SNPs that were not genome-wide significant (P>5e−08), that were in high LD (r2 > 0.1) and that were not in the MTAG GWAS results. The IVs included are shown in supplementary tables 9-12. Although none of the traits past the threshold of multiple testing correction, several findings were reaching the suggestive significant threshold (P < 0.05).

#### Ocular Traits

We chose SNPs for spherical equivalent (12 SNPs), axial length (2 SNPs), IOP (51 SNPs), glaucoma (14 SNPs), AMD (5 SNPs), diabetic retinopathy (2 SNPs) as IV. Interestingly, we found that the smaller spherical equivalent (more myopic) might be causally associated with a greater RAG, although subsequent sensitivity analysis couldn’t support this finding. (IVW: beta=-0.065, P=0.01; MR-Egger beta=0.13, P=0.33; MR-Egger intercept=-0.02, P=0.15) (supplementary table 9). Besides, IVW analysis also suggested that higher IOP tend to accelerate retinal aging (IVW: beta=0.022, P=0.04). Although MR analysis didn’t support the causal effect of AMD on retinal aging, there was a strong heterogeneous effect of the IVs for AMD (Q = 15.52, P= 0.004), which indicated pleiotropy.

#### Modifiable clinical risks

We found that the increasing genetic predisposition to raised glycated hemoglobin levels (8 SNPs) would cause an increase RAG (IVW: beta=0.29, P=0.04; Weighted Median: beta=0.34, P=0.01; Weighted Mode: beta=0.35, P=0.03; MR-Egger intercept=-0.004, P=0.52). For the blood lipid level, the increased HDL level seemed not to be associated with accelerated retinal aging (IVW: beta=0.06, P=0.12) while total cholesterol (IVW: beta=-0.06, P = 0.02), triglycerides (IVW: beta=-0.10, P = 0.03;) and LDL level ((IVW: beta=-0.07, P = 0.01) seem to be causal factors for delaying the senility retina, which is consistent with previous findings about the effect of blood lipid on AMD^43,43^. Blood pressure, blood urea nitrogen levels or eGRF levels were not causally associated with retinal aging (supplementary table 10).

#### Haematological measurements

Since in the PheWAS look-up, we found an association between RAG SNPs and haematological measurements, we tested whether there’s a causal relationship between hemocyte components and retinal aging. Among the tests, mean corpuscular hemoglobin concentration (IVW:beta=-0.13, P = 0.01; Weighted Median: beta=-0.21, P=0.003; Weighted Mode: beta=-0.22, P=0.02;) and basophil percentage of white cells (IVW:beta=-0.13, P = 0.01) were causing a younger retinal appearance, while the inflammation related hemocytes, such as neutrophil percentage of white cells (IVW:beta=0.15, P = 0.002), white blood cell count (IVW:beta=0.10, P = 0.01) and granulocyte count (IVW:beta=0.09, P = 0.036) were causing an older retinal appearance (supplementary table 11).

#### Anthropometric measures and lifestyle

Finally, we investigate the anthropometric measures and lifestyle traits, no causal relationships were identified by MR (supplementary table 12).

## Discussion

In summary, we used different deep learning algorithms to quantify in vivo retinal age across nearly 40,000 human retinal fundus images from two independent study cohorts. Our study has several main findings: 1) Robustness of DL in predicting RAG: although the defining and the deriving of retinal age by deep learning algorithms haven’t been fully established, consistent findings from different studies support the robustness of applying deep learning to quantify the retinal age; 2) Biological relevance of RAG: genetic factors are contributing to the accelerated retinal aging, which explains around 15% of the phenotypic variation; 3) Comprehensive causality: acceleration of retinal aging is a consequence of ophthalmic and whole systemic degeneration, including changing of refractive status, blood biochemistry and hemocytes components etc. Here we would like to discuss these findings in more detail.

### 1. Consistent genetic findings for RAG quantified by different DL suggest the robustness of the DL model

To our knowledge, this is the first study that compared the genetic composition of RAG defined by different DL algorithms and we identified highly overlapped genetic aetiology of RAG between the UK Biobank study and the GoDARTS study. For these two cohorts, despite their discrepancies in the training population and algorithms, many of the genetic findings were replicated, and the correlation of the SNPs was high (Pearson correlation coefficient = 0.82, Figure 1). More importantly, the overall genetic correlation between the two RAGs was as high as 0.67 (P=0.021). These findings partially eliminated our concerns, which is: when there’s no stander or clear definition of what RAG is, will a different model give a totally different trait?

Biological age derived by DL shared common genetic aetiologies, suggesting the information extracted by DL is not artificial. Conventional methods could only extract limited information from these medical data, but DL technology empowers the medical data with more information, such as biological age. As further validation, we compared our findings with Goallec et al’s^20^ findings. In his study, the DL method was applied to different eye images, abdominal MRI, blood test results and lung functions and shared genes were identified among different RAGs as well as different biological ages. The concordance of genetic findings between our defined RAG with a third DL-trained RAG provided further evidence of the reproducibility of DL.

### 2. GWAS findings support the biological relevance of RAG

Firstly, we found that RAG calculated by different DL is replicable, which built up the basis for looking for the biological relevance of the trait. Then, by using a hypothesis-free genome-wide association analysis, we explored the genetic factors of RAG. We used MTAG to merge two GWAS results. MTAG is a recently developed meta-analysis method that is known for its robust character against sample overlapping and less concordance in traits^26^. The MTAG boost the power of GWAS and identified new loci for retinal aging. According to our findings, we estimate that genetic factors can explain around 15% of the variation of the trait. In other words, there are genetic factors that determine the ‘younger’ or ‘order’ appearance of the retina compared to the average age-matched population.

The GWASs and gene-based enrichment analyses performed on the RAG traits highlighted genetic variants or genes associated with retinal function or even retinal aging mentioned above by Goallec et.al.^20^ (Figure 5 a). *SH3YL1* (SH3 And SYLF Domain Containing 1), is the top GWAS signal in our study. and a recent study unveils that *SH3YL1* is a gene regulating the retina vessel density. Together with *OCA2* (oculo-cutaneous albinism 2), which is required for melanosome maturation and involved in post-embryonic morphogenesis in the eye of fish^47^, is a newly identified gene for retinal vessel fractal dimention^12^; These findings may suggest that the retinal vessel morphology may alter the blood perfusion hence causing the degeneration. Another set of genes - *PLEKHA1/ARMS2/HTRA1*- located on chromosome 10, were identified in the GWAS for the GoDARTS population only, and are genes for AMD^48,48^. Although not replicated in our GWAS on the UKB population, which is probably due to the relatively ‘healthier’ images being trained in the UKB population, Goallec et.al’s^20^. GWAS for RAG on eye image supported our findings. This indicates that AMD genes are driving retinal aging. Among the other replicated RAG genes, *MACF1* (microtubule-actin crosslinking factor 1) is one of the most abundant proteins of the photoreceptor proteome^51^, which also seemed to be involved in brain aging^52^, brain aging diseases like Parkinson^53^ and aging-related bone loss^54^; *FAM150B/ACP1* (platelet-derived growth factor receptor signalling) is involved in myopia development^55^. *PPL, ST3GAL6-AS1*, were found for accelerating the aging process of the lungs or abdominal (Figure 5b). While *CCDC25* is related to Alzheimer’s disease^56^. The partially overlapped aging genes among different organs might indicate although there is a uniform chronological age there might be a heterogeneous biological age for different tissue/organs.

Several newly identified RAG genes also demonstrated effects on age-related phenotypes. Seventeen genes were not reported for RAG previously. *TMCC2* (Transmembrane And Coiled-Coil Domains Protein 2): TMCC2 binds with APOE^57^, which is a well known gene for longevity. *DSTYK* (dual Serine/Threonine and Tyrosine protein kinase) is involved in the skin aging process^58^; *SCARA3* (Scavenger receptor class A, member 3) is highly expressed in autosomal recessive retinitis pigmentosa eyes^59^; *KCNQ1DN* is hypermethylated upon aging and was used as a biomarker to predict epigenetic-aging^60^.

In summary, by performing a GWAS on RAG, we identified the potential genetic contributions to accelerated retinal aging which support its biological relevance. This further support that RAG might be a good biomarker for aging as it also reflects the systemic aging process.

### 3. Both ophthalmic and systemic factors accelerate retinal aging

Identifying the causal factors for RAG is important. As a newly emerged concept, ‘accelerated retinal age’ are gradually getting popular, but this artificially defined trait remains the ‘black box’ nature inherited from DL. On one hand, exploring its potential in the future disease prediction area could maximize its clinical application; while on the other hand, investigating its biological relevance or even its biological causal mechanisms could open the ‘black box’ and unveil the etiologies of acceleration of retinal degeneration. By using the technique of Mendelian randomization, we assess the biological causal factors for RAG from the genetic basis for the first time. In our MR study, although none of the traits passed Bonferroni correction, we still value the findings suggested by MR. In general, the 95% CIs of the IV estimates were wider compared to those of the conventional regression, hence a significant finding via IV analysis (P < 0.05) usually indicating strong phenotype associations. Besides, we applied a series of sensitivity analyses for each trait to increase the robustness of our findings.^61,62^

According to our MR findings, we notice that the IVW method suggests that myopia might accelerate retinal aging. This is contradictory to the observational study that a positivist association was observed between SE and chronological age. This might be due to the following reasons: 1) RAG reflects the biological function of the retina in a better way compared to the chronological age. Myopia, especially high myopia, would cause thinning of the retinal neuron layer and increase the risk of retinal detachment and AMD^63^, which are all causes of retinal degeneration and vision loss. 2) RAG represent the retinal status other than the overall ocular refractive situation. The positivist association between SE and chronological age is caused by presbyopia -a consequence of the loss of the ability for the eye to accommodate to focus on nearby objects. This is mainly due to the loss function of the lens but not the retina, RAG might not capture this information.

For the modifiable clinical risk factors, we notice that HbA1c is a strong causal factor for RAG. Elevated HbA1c is the consequence of long-term expose to hyperglycemia, and is the hallmark of T2D. Glycosylation and reactive oxygen are induced during the hyperglycemia exposure, which would cause vascular endothelial dysfunction or death, leading to a leakage of the micro vessels^64,65^. The clinical manifestations are featured as micro-aneurysm, exudes, or even neovasularization^66^, which influence the function of the retina and might be the marker of retinal aging. Besides, our MR results suggest elevated LDL and cholesterol are associated with a younger appearance of the retina. Different from cardiovascular disease, many MR studies also reported a risk role of HDL and protective roles of LDL/cholesterol/TG on AMD ^43,67,68^. These findings from epidemiologic studies may neglect the competing risk of death^69^. By far, no clear mechanism can explain this phenomenon. According to previous studies, during the formation of HDL and the lipid transportation process within the retina, lipoprotein was accumulated and acts as a barrier for lipid transport through an aging retina, leading to the formation of drusen, the marker of AMD^45^. Meanwhile, HDL may contain essential complement components, which are involving the major pathway in AMD pathogenesis ^70^. Our findings highlight the value of studying the biological age of an organ, since different organs are susceptible to different risk factors.

We also notice that increased hemoglobin concentration is causally associated with a younger appearance of the retina while increased white cell counts and neutrophil percentage accelerate retinal aging. There is rich blood perfusion into the retinal choroid and the retinal neuron layer is nourished by the ocular vessel system. Anemia is featured as low hemoglobin concentration and leads to low oxygen-carrying capacity in the blood. Anemia causes retinal hypoxia, which can lead to infarction of the nerve fibre layer and clinically manifests as cotton wool spots. Retinopathy is a frequent finding in anemic and thrombocytopenic patients and our findings confirmed this findings^71^. White blood cells and neutrophils are the inflammatory markers. During the inflammation reaction process, reactive oxygen species are generated, while the oxidative stress is associated with a wide range of retinal diseases such as AMD ^72^, diabetic retinopathy^73^ and some rare retinal diseases such as Leber Hereditary Optic Neuropathy^74^. For systemic diseases, with the aging process, granulocyte counts were increased which is also related to Parkinson’s disease ^75^. These findings indicate that the hemocyte components are also related to retinal aging and RAG also inversely unveils the systemic aging process.

### 4. Limitations

The comparability of different DL methods needs to be assessed systematically by comparing each other within the same population. During the analysis, we found that both the UKB model or GoDARTS model tend to shrink the distribution of the predicted retinal age. E.g., For people with younger chronological age, their predicate age tends to be older and vice verses. To overcome this problem, in our GWAS analysis, we used the rank transformation to overcome this problem partially. Lastly, our study is unable to describe the environmental effect of retinal aging, hence epidemiology studies are needed.

### 5. Conclusion

In summary, we discovered and replicated the genomic determinants of retinal biological age estimated by DL applied to fundus images by GWAS. We also explored the potential causalities through a series of MR analyses. Our findings have several contributions: 1) the concordance findings between two independent cohorts reassumed robustness of DL in predicting retinal imaging-based ages; 2) GWA results and following bioinformatic analyses provided the genetic etiology of retinal aging; 3) MR findings indicated that the retina is particularly vulnerable to risk factors such as elevated glycated hemoglobin, inflammation factors or anemia, hence accelerated retinal age might reflect the corresponding systemic conditions. Our findings expanded the current knowledge of the biological implications of retinal aging as well as provided potential therapeutic targets for the general aging process.

## Methods

### UK Biobank cohort

UK Biobank recruited more than 500,000 UK residents aged between 37-73 years during 2006 and 2010. Participants were invited to complete comprehensive healthcare questionnaires, provide detailed physical measurements, and provide biological samples. Other laboratory tests, medical images or health-related events were ascertained via data linkage to hospital admission records and mortality registry. The majority of individuals who consented were also genotyped. The baseline examination resulted in the collection of 68,151 paired colour retinal photographs and was used in this study. Ethical approval was obtained from the National Health Research Ethics Service (Ref 11/NW/0382) and all participants provided (digital) written informed consent. Details of the study design and data composition can be found elsewhere.^21^

### GoDARTS cohort

Participants from the Genetics of Diabetes Audit and Research in Tayside Scotland (GoDARTS) were tested in the replication phase. The GoDARTS type 2 diabetes (T2D) case-control study population was recruited from the Tayside region of Scotland between the years 1997 and 2007, for each participant, the longitudinal data on biochemical investigations, prescriptions, clinical events were assembled through data linkage to their electronic medical records either from hospital admission, or primary health care. In total, more than 20000 participants with medical records for maximum 30 years of follow up were assembled. A subset of these participants had also consented for genetic analysis (genome-wide genotyping). GoDARTS has previously been described fully^22^. In this study we used diabetes retinal screening images obtained from the Scottishe Diabetes retinal Screening Service which were all centrally collected since 2006. More details can be found at http://diabetesgenetics.dundee.ac.uk/.

The GoDARTS study has been approved by Tayside Committee on Medical Research Ethics, and informed consent was obtained from all patients (REC reference 053/04).

### Deep Learning estimation of retinal age and retinal age gap

For details of the training and verification process of the DL model for age prediction in UK Biobank please refer to Zhu et.al^18^. Briefly, within the 46,969 UK biobank participants whose retinal fundus images were available, 11,052 participants who reported no previous disease were used to train the DL model for age prediction. Of the remaining 35,917 participants, images from the right eye were prioritized for the prediction of retinal age.

In the GoDARTS study, only patients with T2D had retinal images. In total, 8,570 individuals had retinal images at the baseline. During the follow up yearly screening, a total number of 102,082 retinal images from both eyes were collected. These images were used from training to derive the retinal age, and the baseline retinal age was used for analysis in this study. Details of the DL process have been provided in Supplementary method 1.

For both the UK Biobank and GoDARTS cohort, we defined the difference between retinal age predicted by the DL model and chronological age as the RAG. A positive RAG, indicates predicted age being older than chronological age. The rank-based inverse normal transformation (INT) was applied to the RAG in the main study in order to normalize RAG. In the sensitivity analysis, RAG was analyzed as a continuous trait or binary trait - greater than 3.55 years (mean absolute error) as the ‘older group’ and less than -3.55 years as the ‘younger group’.

### Genotyping, imputation and quality control

For the UK Biobank, genome-wide genotype data were available for 488,377 participants. For the GoDARTS participants, genome-wide genotyping was available for 25,409 patients with diabetes. Details of the genotyping and imputation process are in supplementary method 2.

In general, routine genetic quality control was applied before the analysis: SNPs with call rate < 95%, imputation quality score < 0.8, or with minor allele frequency (MAF) less than 1%, or those that failed Hardy–Weinberg tests (P > 10e-06) were removed; SNPs on the sex chromosomes and mitochondrial were also excluded from analyses. For sample QC, individuals who don’t have the retinal image, were not European ancestry, with genotyping call rate < 95%, or showed a mismatch between phenotypic and genotypic gender, or demonstrate relatedness with others (UK Biobank: third degree relatives or closer; GoDARTS IBD > 0.8) were removed.

### GWAS and meta-analysis

In stage 1, we performed GWAS of RAG in the UK Biobank population. We used mixed models (BOLT-LMM v2.4^23^) and adjusted for sex, age and the first 5 principal components. In stage 2, for the RAG GWAS in GoDARTS, we took use PLINK 2.0 software^24^ for the GWAS analysis. However because GoDARTS genotyping has been done on several different arrays this was performed separately for each separate genotype data set. To combine the results within GoDARTS study, we meta-analyzed the GWAS summary statistics using the inverse variance weighted method (METAL software^25^).

Subsequently, in stage 3, cross validation was performed to compare the GWAS results between the two cohorts. Prior to further meta-analysis, the genetic correlation between the two GWASs were evaluated in order to test whether there was shared genetic architecture between them. MTAG (multi-trait analyses of GWAS) ^26^ was run to combine the summary statistics between the UK Biobank cohort and GoDARTS cohort. By incorporating information from other genetic correlated traits, MTAG leverages the common genetic information and boosts the power of the trait of interest. Figure 1 illustrated the overall study design.

### Enrichment analysis

We conducted gene-based and pathway analysis in MAGMA, as implemented in FUMA (version 1.3.1)^27,28^. In general, in the annotation step, a 5kb upstream and downstream window was applied to the gene location information and then gene based analysis was performed. A total 10894 predefined gene sets from KEGG, Reactome, BioCarta and Gene Ontology (GO) terms were used for pathway enrichment analysis^29,29^, and Bonferroni corrected P values were used to define the significance threshold.

### SNP annotation and phenome-wide association analysis (PheWAS)

To identify independent genome-wide significant SNPs, we applied both clumping (r2 < 0.1 within 250kb) and joint and conditional analysis (cojo-GCTA) to the summary statistics from MTAG analysis^33^. For the significant SNPs, variants annotation was based on the Genome Reference Consortium Human genome build 37 derived from the University of California Santa Cruz (UCSC) Genome resource^34^. In silico functional annotation was performed by FUMA^28^ which includes the annotation databases such as RegulomeDB^35^, HaploReg v1^36^. Expression quantitative trait loci (eQTL) analysis was performed using the EyeGEx database^37^ and chromatin interaction mapping was further analyzed by using Hi-C^38^.

To further investigate the cross-trait effects of the RAG SNPs, in PhenoScanner ^39^ and GWAS catalog^40^, We retrieved all association results for SNPs in high LD (r^2^ = 0.8, European ancestry) that were genome-wide significant at P<5e-08. For further explanation and exploration, PheWAS were carried out for each individual SNP by looking up their association with other disease or lifestyle traits in the Gene Atlas catalog (http://geneatlas.roslin.ed.ac.uk).

### Comparison with other biological age genes

As further validation, genes reported from GWAS of ‘eye ages’, ‘lung age’, ‘abdominal age’ or ‘blood age’ were compared with genes for RAG by Venn-diagram. The tissue age genes were reported by Goallec et.al (https://www.multidimensionality-of-aging.net/).

### Genetic correlation

LD Score regression implemented in LDSC^41^ (https://github.com/bulik/ldsc) was used to estimate the SNP heritability (h2) of the RAG. To further investigate the extent to which the genetic variance was shared between accelerated retinal age and other relevant biological traits, genetic correlations were estimated for traits with adequate heritability (h2 > 0.1). These biological traits included ophthalmic traits, anthropometric traits, blood pressure, lipid, glycated hemoglobin, kidney function, smoking, cardiovascular diseases, Alzheimer’s disease, diabetes and longevity. Details of the summary data can be found in the data availability section and supplementary table 8. The summary statistics were downloaded through GWAS catalog links^40^.

### Mendelian Randomization

Finally, to explore the potential causalities of the RAG, Mendelian Randomization analysis was performed using Two-Sample MR in R package^42^. These traits include ocular features-axial length, refractive error, intraocular pressure, glaucoma, age-related macular degeneration (AMD) and diabetic retinopathy; anthropomorphic and lifestyle traits - height, BMI, smoking, alcohol consumption and sleep duration; biological traits-blood pressure, blood lipids, blood glucose and hemocytes. Details of the data can be found in supplementary tables 9-12. The instrumental variables for each trait were taken from SNPs reported in GWAS catalog, with certain QC processes: removing ambiguous SNPs, removing miss match SNPs, paradomic SNPs and SNPs in high LD (r2>0.1).

## Author contributions

Study concept and design: Huang Y, He MG, Doney ASF, Zhu ZT; Generation of Retinal age: Syed MG, Trucco E, Mookiah MRK; Access to raw data: Huang Y, Wang H, Doney ASF, Zhu ZT, Srinivasan S, Wang W, Chen RY; Acquisition, analysis, or interpretation: All authors; Drafting of the manuscript: Huang Y, Doney ASF, Zhu ZT; Critical revision of the manuscript for important intellectual content: Doney ASF, Wang H, Shang XW, Yu HH, Hu YJ, Zhang XL, Zhang XY Tang SL; Statistical analysis: Huang Y, Li C; Obtained funding: He MG, Doney ASF; Administrative, technical, or material support: Liu J, Liu SM; Study supervision: Zhu ZT, Doney ASF, He MG.

## Data Availability

Data are available in a public, open access repository (https://www.ukbiobank.ac.uk/). The GWAS summary statistics used in this study for genetic correlation or MRanalysis are available via GWAS Catalog, under study accession identifiers GCST007461, GCST006368, GCST001032, GCST002647, GCST007982, GCST007327, GCST008062, GCST002221, GCST006630, GCST008059, GCST007954, GCST002223, GCST002222, GCST006629, GCST006624, GCST002216, GCST001884, GCST002115, GCST006065, GCST005580, GCST006424, GCST006289, GCST005195, GCST006906, GCST007511, GCST006867, GCST006414, GCST008064, GCST008598, GCST009890, GCST009413, GCST90011766, GCST009404, GCST009411, GCST009412, GCST009414 or PMID:29227965. Summary statistics of body height and BMI were acquired from GIANT consortium (https://portals.broadinstitute.org/collaboration/giant/index.php/GIANT_consortium_data_files). The data and code used in this current study are available from the corresponding authors on a reasonable request.

## Acknowledgement

This research was conducted using the UK Biobank and the GoDARTS resource. We thank the participants of the UK Biobank and participants from the University of Dundee.

## Funding

Huang Y receives support from the Research Foundation of Medical Science and Technology of Guangdong Province, China (A2022323); NSFC Incubation Project of Guangdong Provincial People’s Hospital, China (KY0120220051) and Science and Technology Program of Guangzhou, China (202002020049); Zhu ZT receives support from the National Natural Science Foundation of China (81870663, 82171075), the Outstanding Young Talent Trainee Program of Guangdong Provincial People’s Hospital (KJ012019087), Guangdong Provincial People’s Hospital Scientific Research Funds for Leading Medical Talents and Distinguished Young Scholars in Guangdong Province (KJ012019457), Talent Introduction Fund of Guangdong Provincial People’s Hospital (Y012018145). He MG receives support from the University of Melbourne at Research Accelerator Program and the CERA Foundation. The Centre for Eye Research Australia receives Operational Infrastructure Support from the Victorian State Government.

## Data and code availability

Data are available in a public, open access repository (https://www.ukbiobank.ac.uk/). The GWAS summary statistics used in this study for genetic correlation or MRanalysis are available via GWAS Catalog, under study accession identifiers GCST007461, GCST006368, GCST001032, GCST002647, GCST007982, GCST007327, GCST008062, GCST002221, GCST006630, GCST008059, GCST007954, GCST002223, GCST002222, GCST006629, GCST006624, GCST002216, GCST001884, GCST002115, GCST006065, GCST005580, GCST006424, GCST006289, GCST005195, GCST006906, GCST007511, GCST006867, GCST006414, GCST008064, GCST008598, GCST009890, GCST009413, GCST90011766, GCST009404, GCST009411, GCST009412, GCST009414 or PMID:29227965. Summary statistics of body height and BMI were acquired from GIANT consortium (https://portals.broadinstitute.org/collaboration/giant/index.php/GIANT_consortium_ data_files). The data and code used in this current study are available from the corresponding authors on a reasonable request.

## Declarations

### Conflict of interest

None of the authors has any conflicts of interest to disclose.

### Ethics approval and consent to participate

This study was approved by the National Information Governance Board for Health and Social Care and the NHS Northwest Multicenter Research Ethics Committee (11/NW/0382) and the Biobank consortium (application number 62489). Since de-identified data in a public dataset was used, the Medical Research Ethics Committee of Guangdong Provincial People’s Hospital waived the requirements to obtain ethical approval.

## Notes

### Competing Interest Statement

The authors have declared no competing interest.

