## Supplementary Figures for "Genomic Determinants of Biological Age Estimated By Deep Learning Applied to Retinal Images"

| 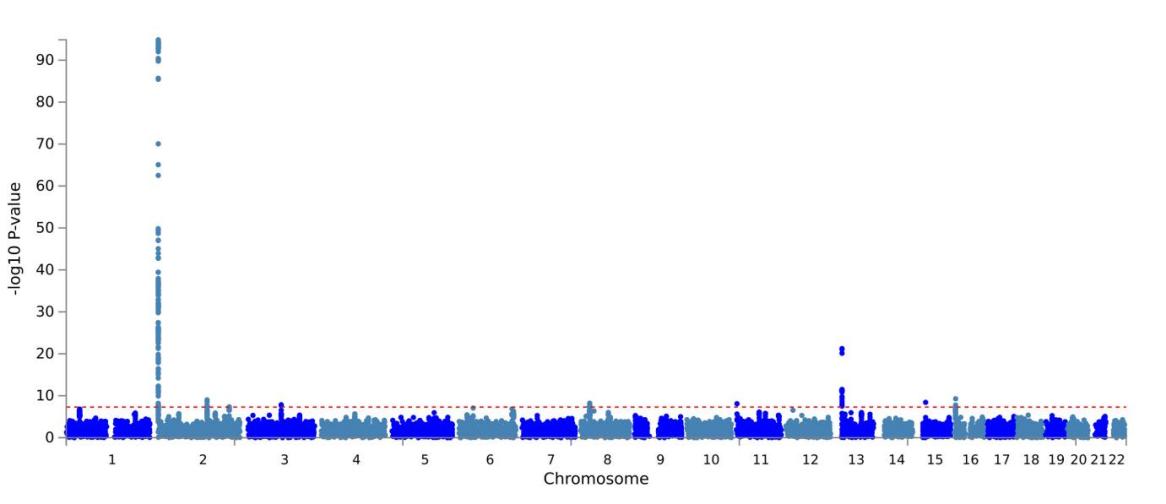 | A |
| --- | --- |
| 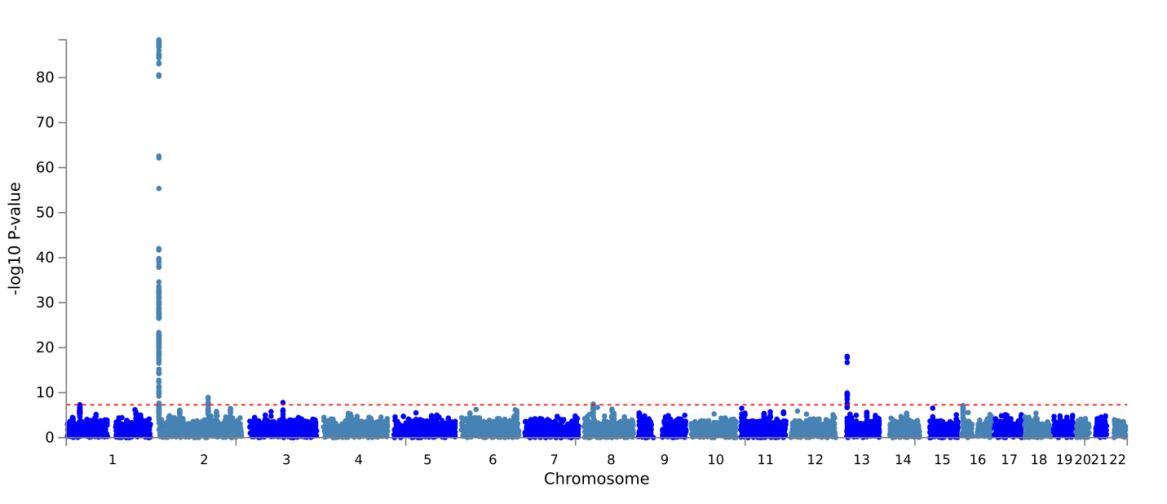 | B |
| 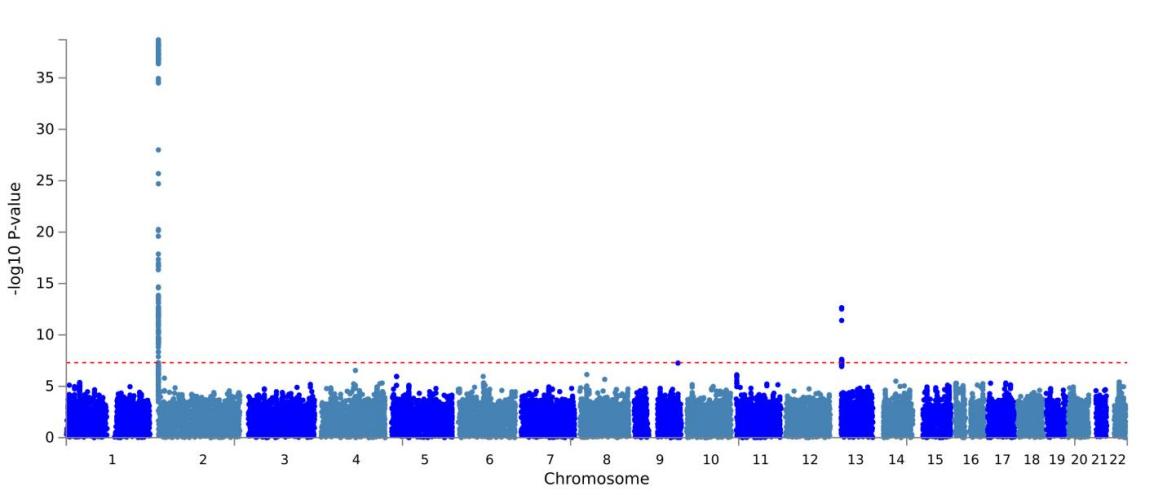 | C |

**Supplementary Figure 1.** Manhattan plot of GWAS results of RAG in the UKB population. A) IVW transformed RAG (Mix model); B) Non-transformed RAG (Linear model); C) logistic model by taking RAG < -3.35 as control and RAG > 3.35 as cases. The x-axis shows the chromosomes where each SNP is located, and the y-axis shows −log10 P-value of the association . The dash horizontal line shows the genome-wide significant threshold (P-value = 5e-8 ).

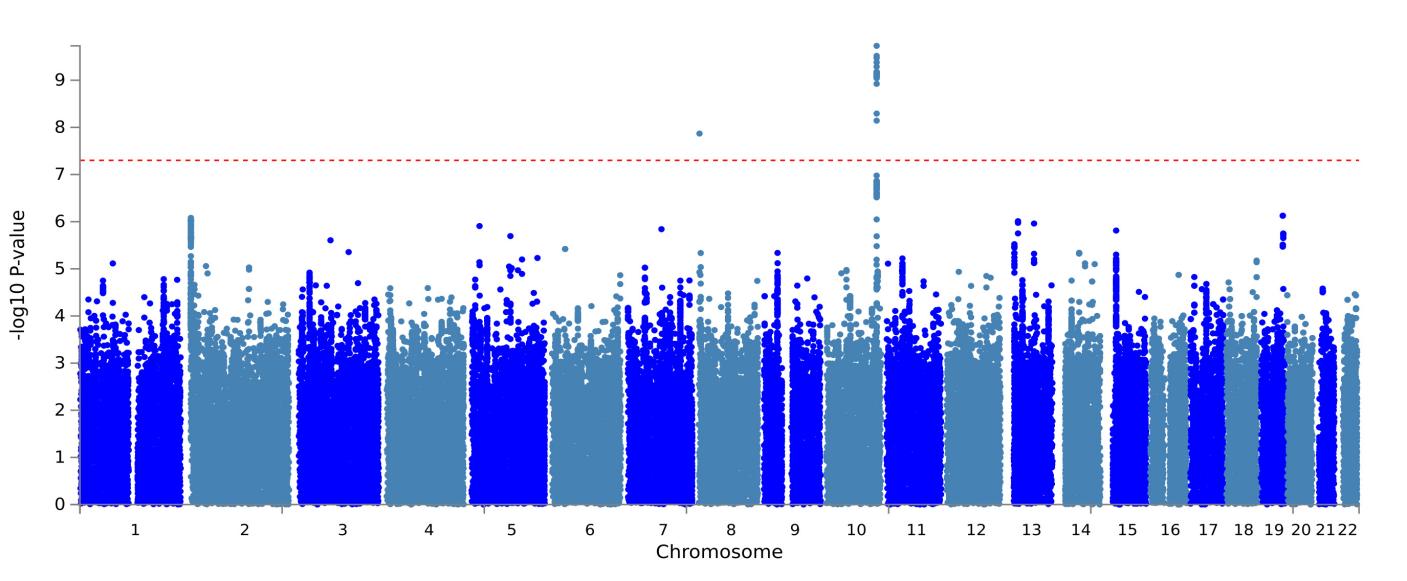

**Supplementary Figure 2.** Manhattan plot of GWAS results of RAG in the GoDARTS population. Each dot represents a SNP, the x-axis shows the chromosomes where each SNP is located, and the y-axis shows −log10 P-value of the association of each SNP with RAG in the stage 2 GoDARTS replication analysis. The dash horizontal line shows the genome-wide significant threshold (P-value = 5e-8 ).

| 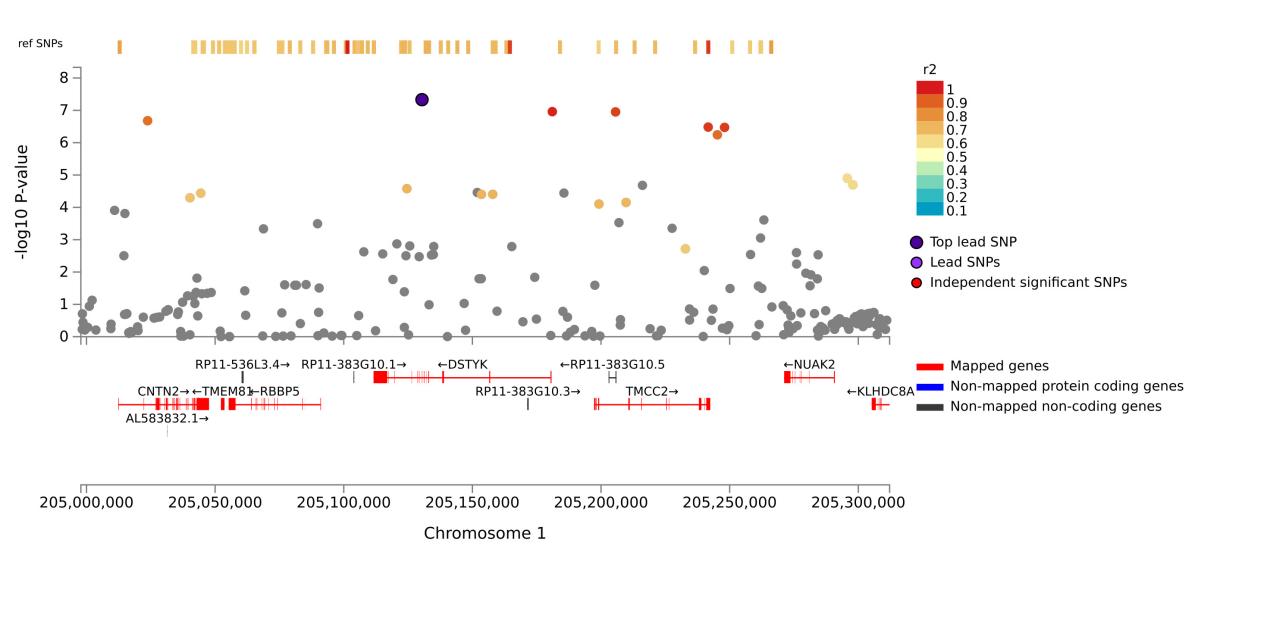 | A  rs3851294 |
| --- | --- |
| 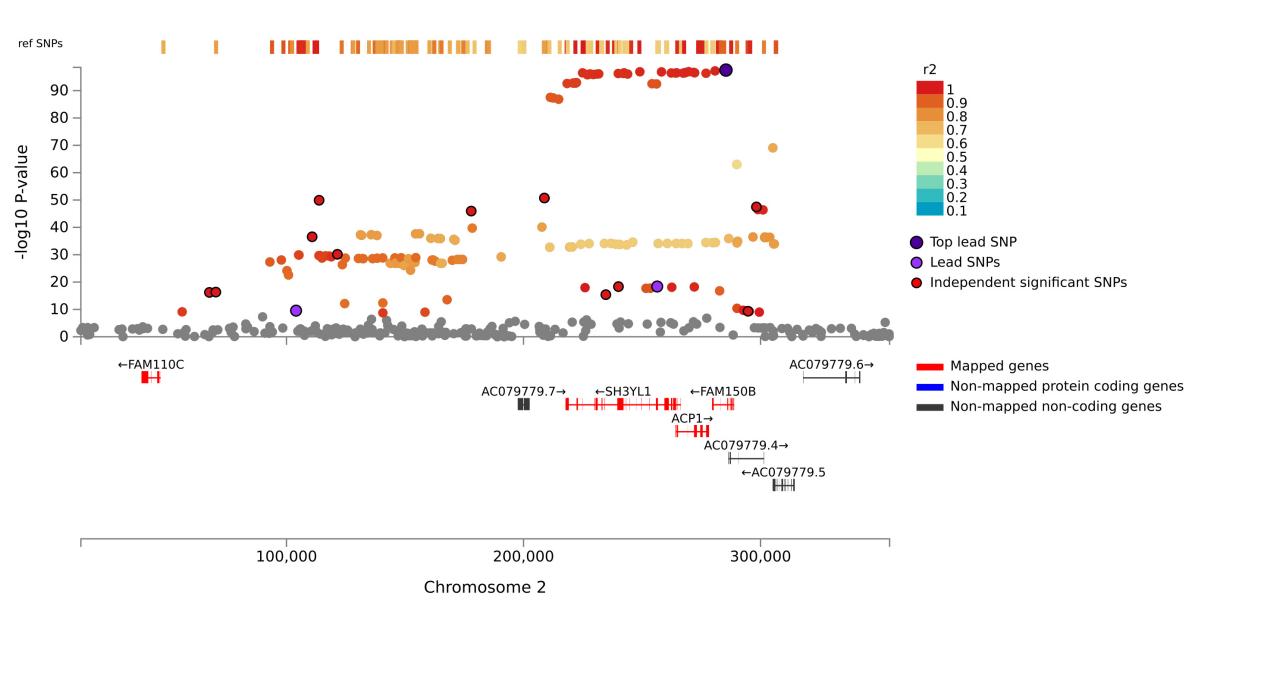 | B  rs60149603 |
| 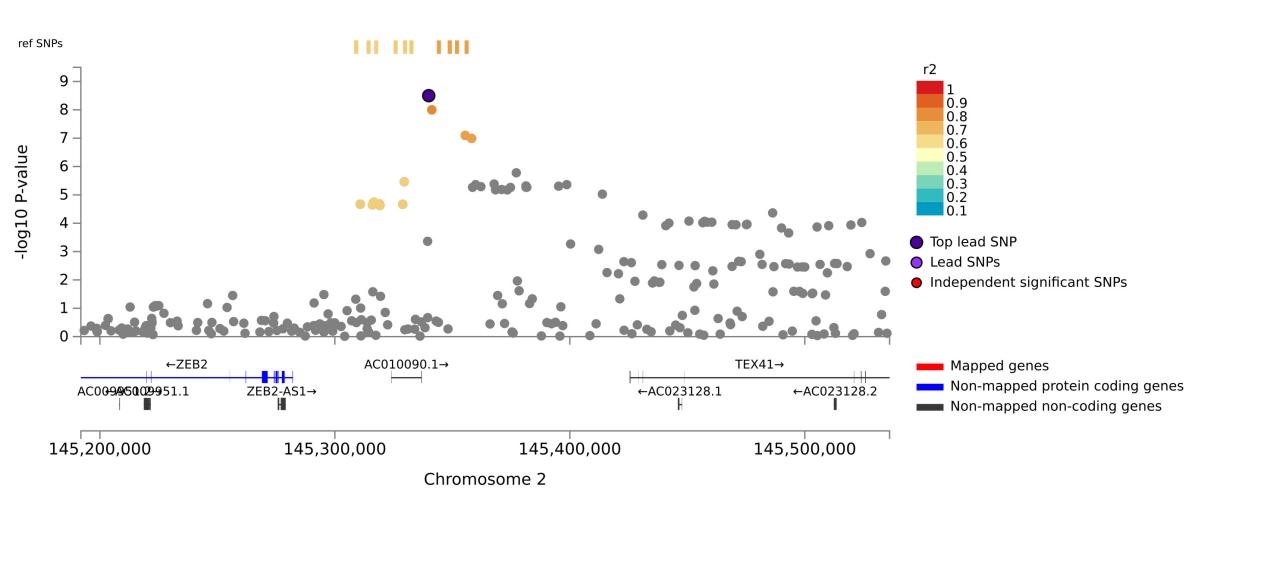 | C  rs4143008 |
| 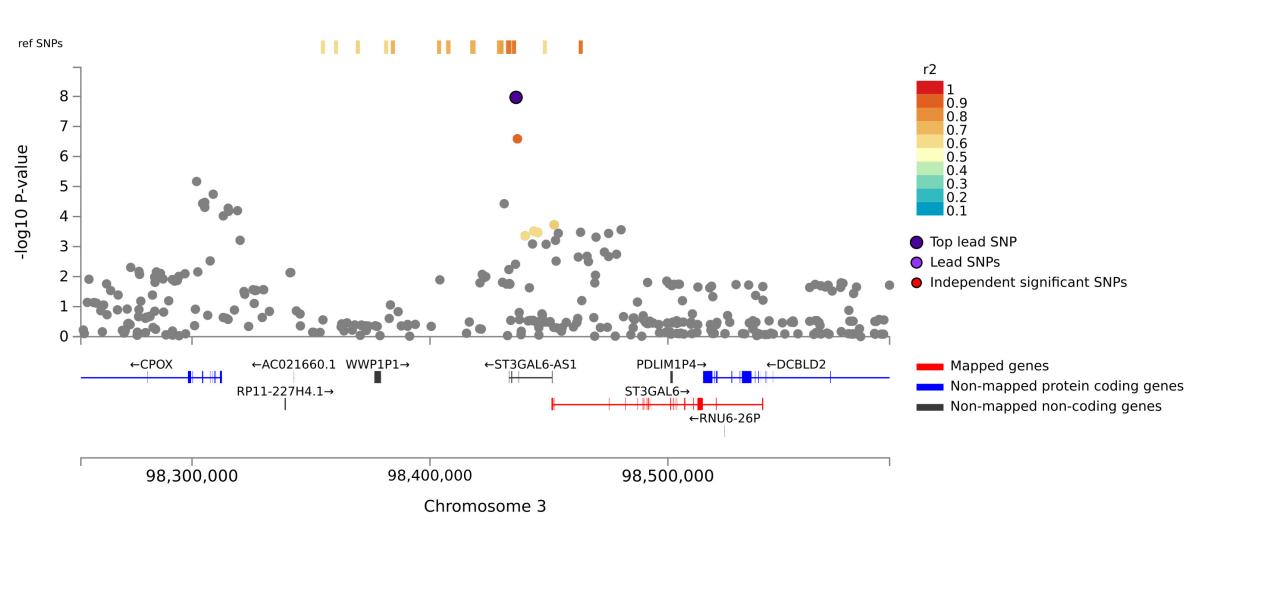 | D  rs12635955 |
| 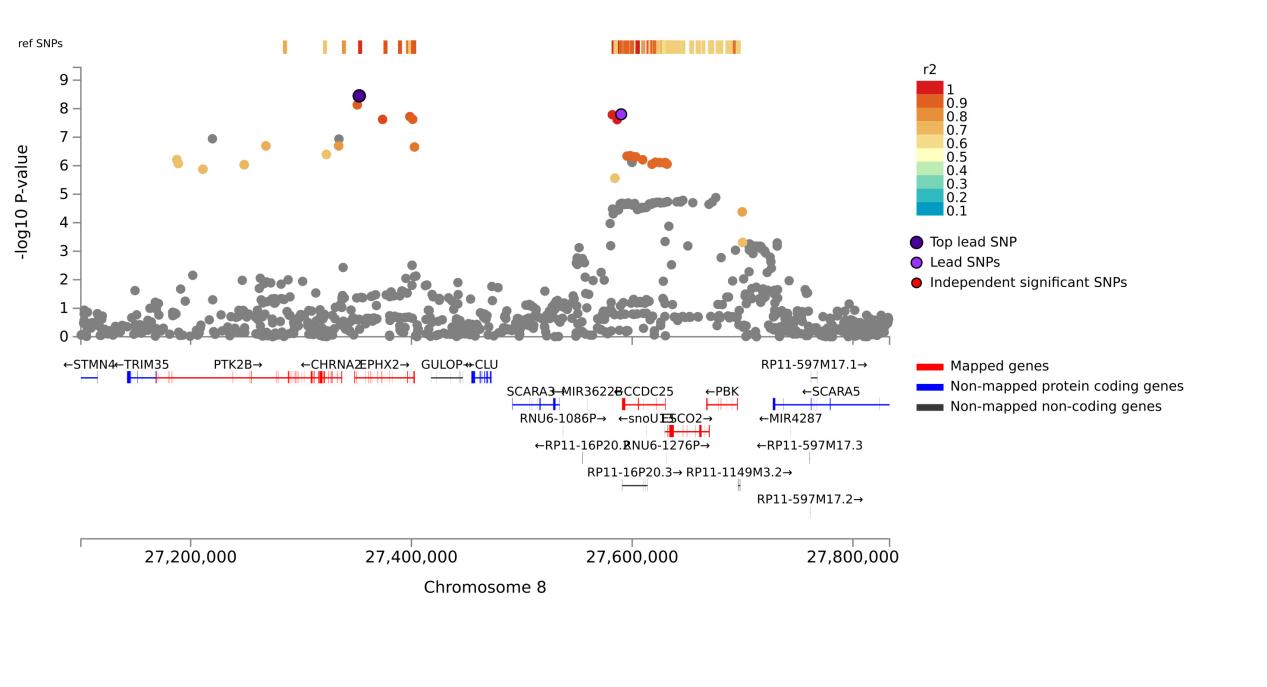 | E  rs2322721 |
| 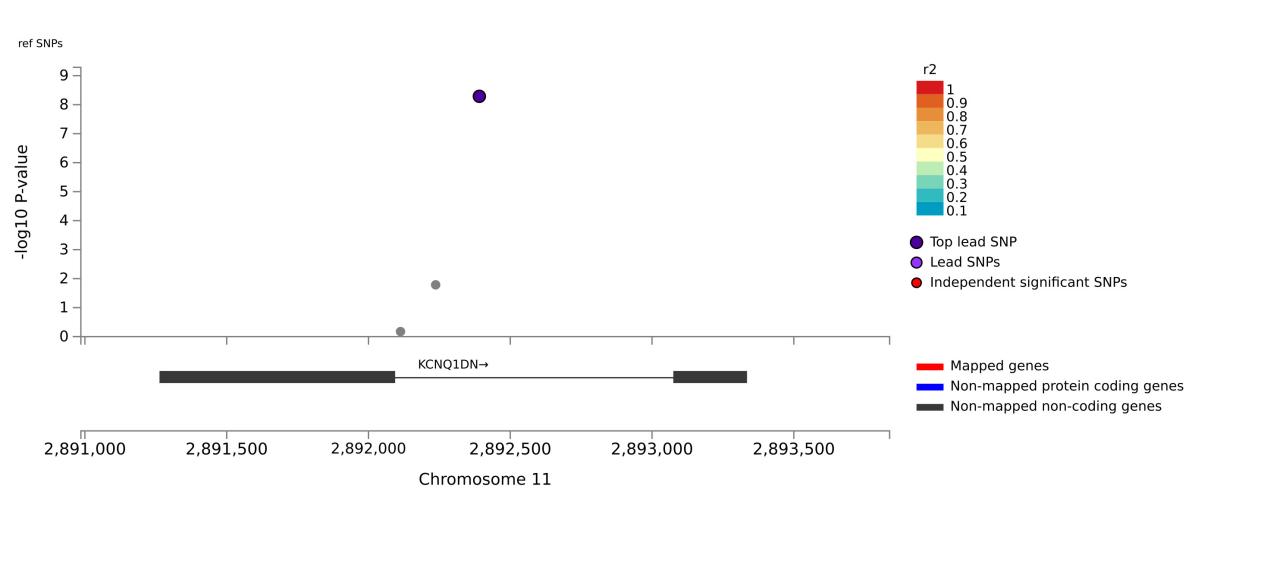 | F  rs4930021 |
| 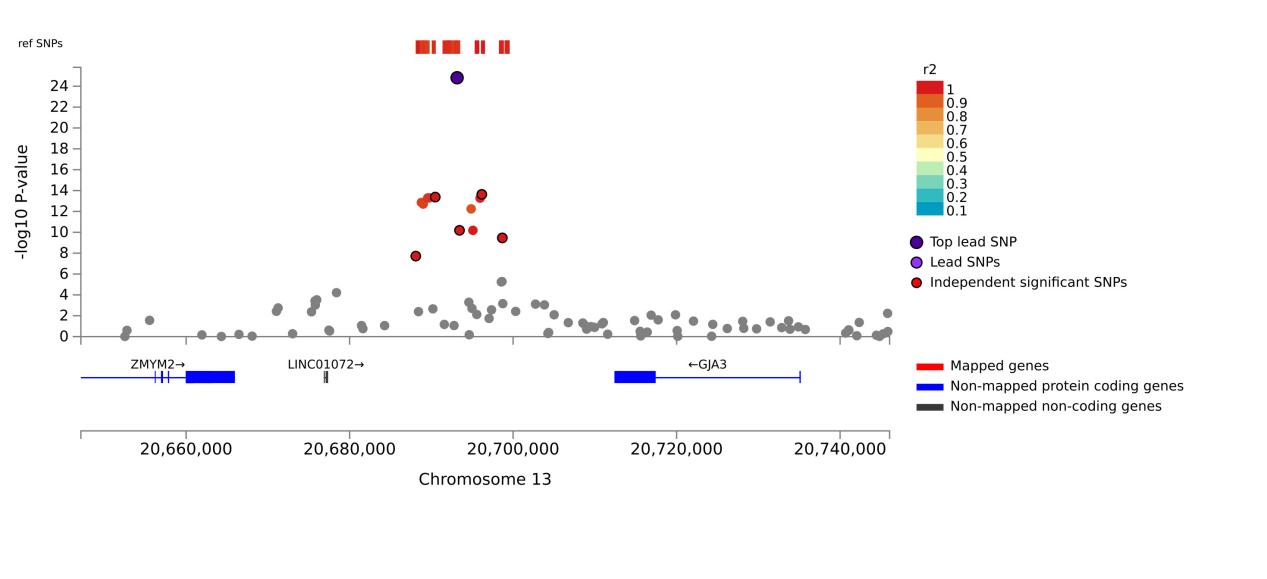 | G  rs8001273 |
| 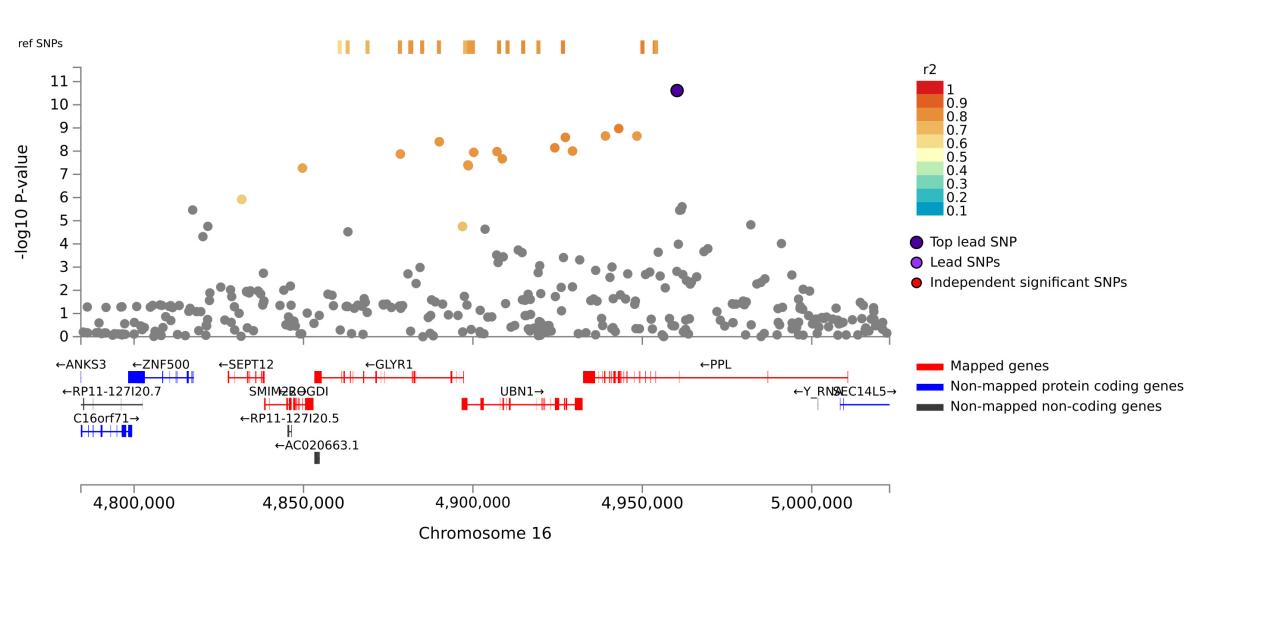 | H  rs8051560 |

**Supplementary Figure 3. Locus Zoom plot for the leading genomic regions of the MTAG results.** From A-H were Locus Zoom plot for the top SNPs located on chromosome 1 (A),chromosome 2 (B), 2 (C), 3(D), 8 (E), 11 (F), 13 (G),16 (H) respectively.

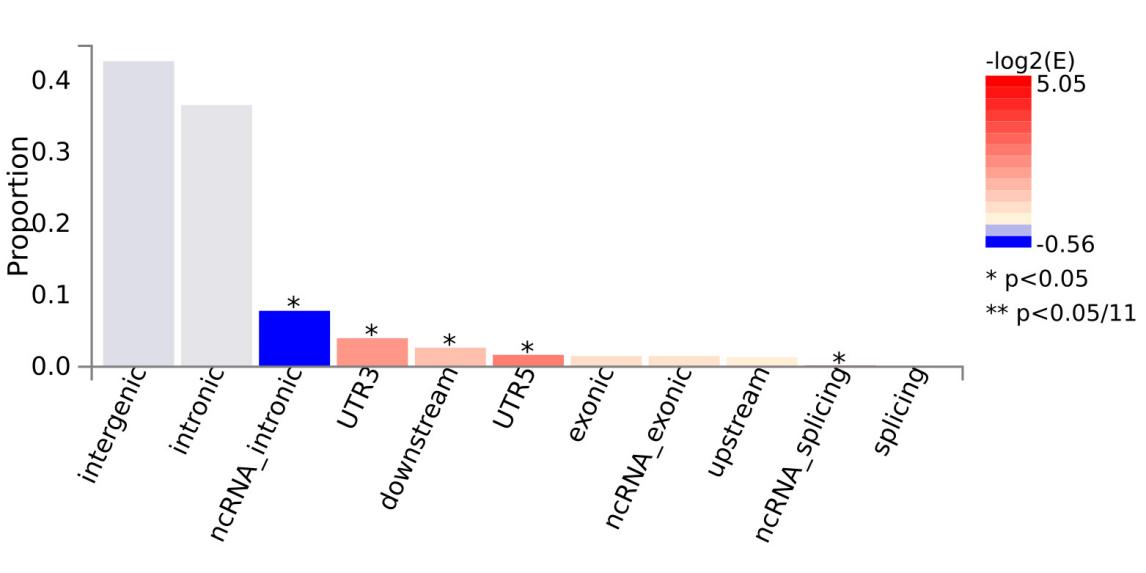

**Supplementary Figure 4. Histogram of the functional consequences of RAG SNPs on genes.**

| 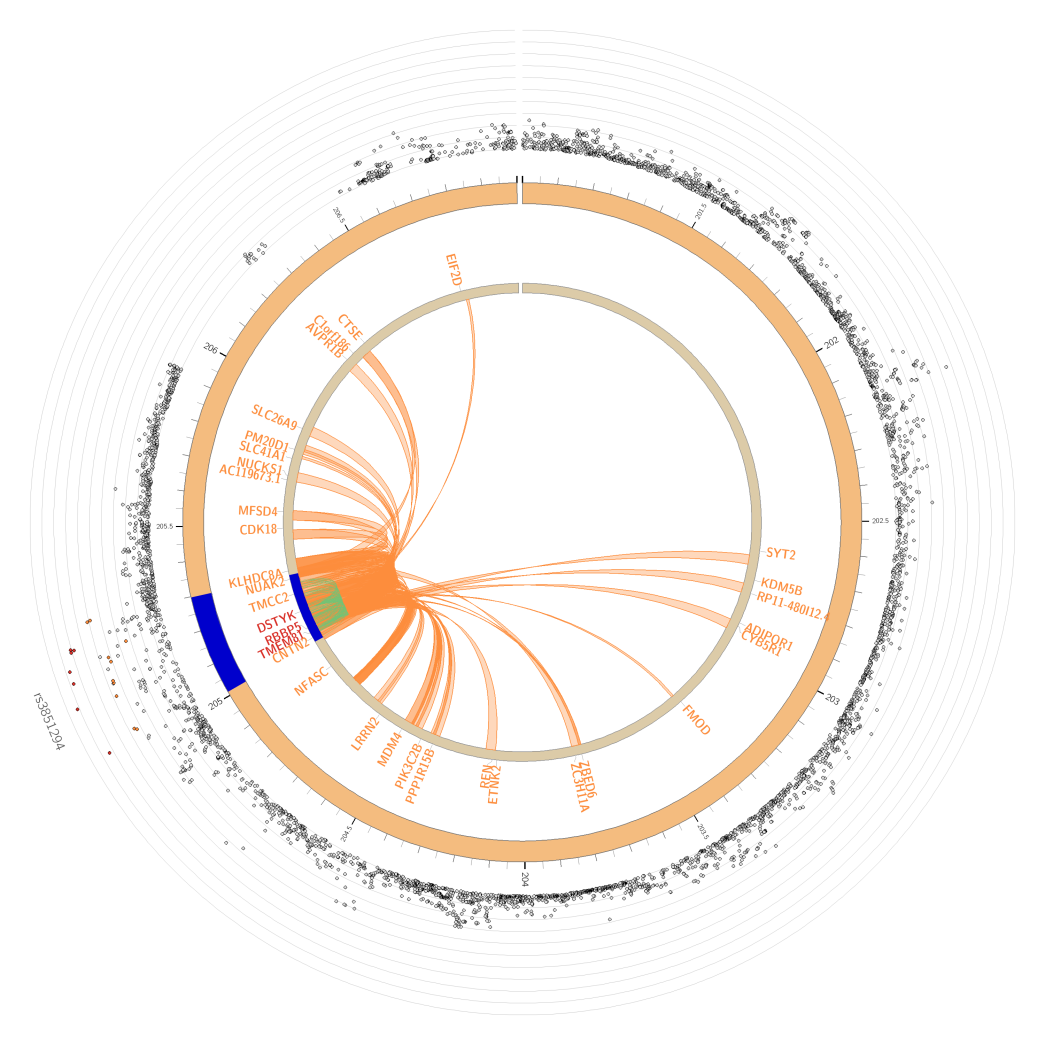 | 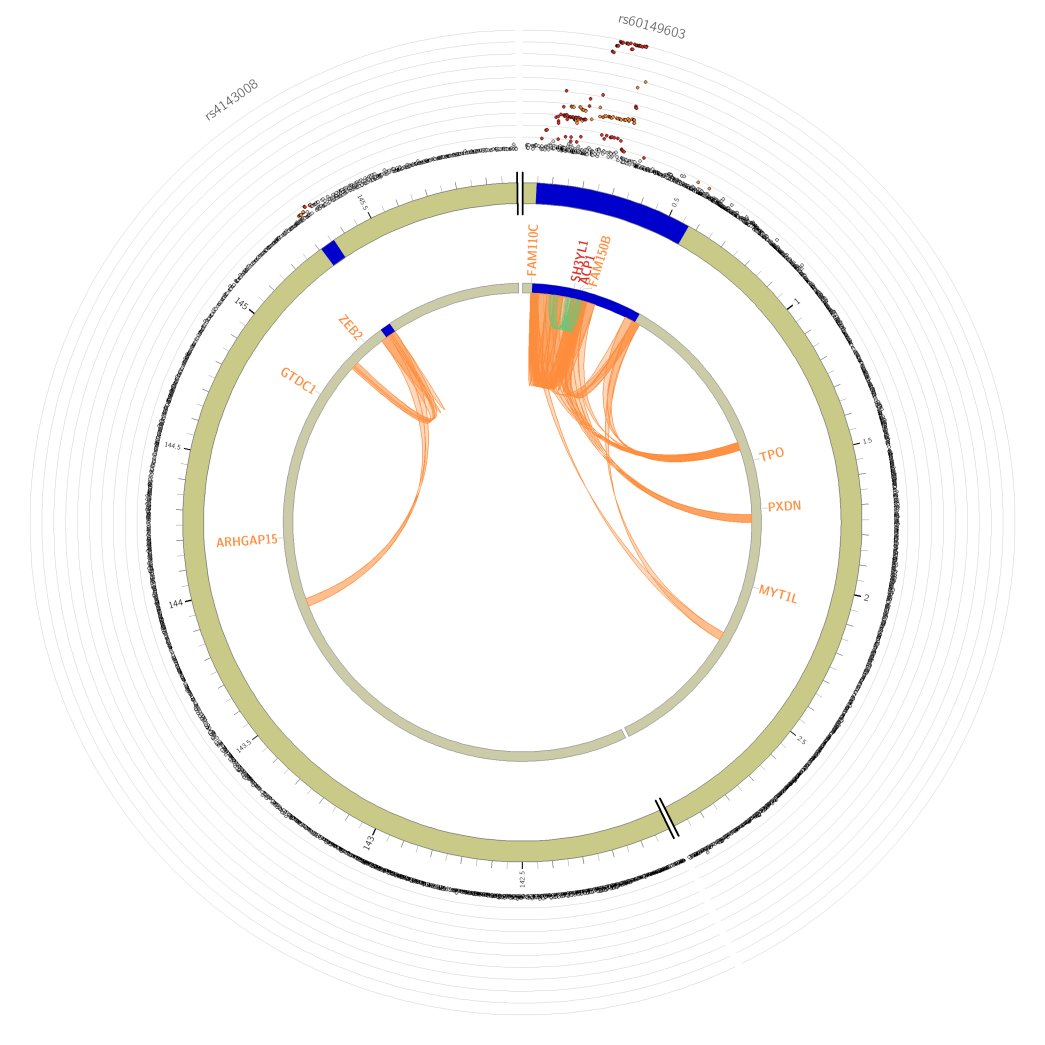 |
| --- | --- |
| A) Chromosome 1 | B) Chromosome 2 |
| 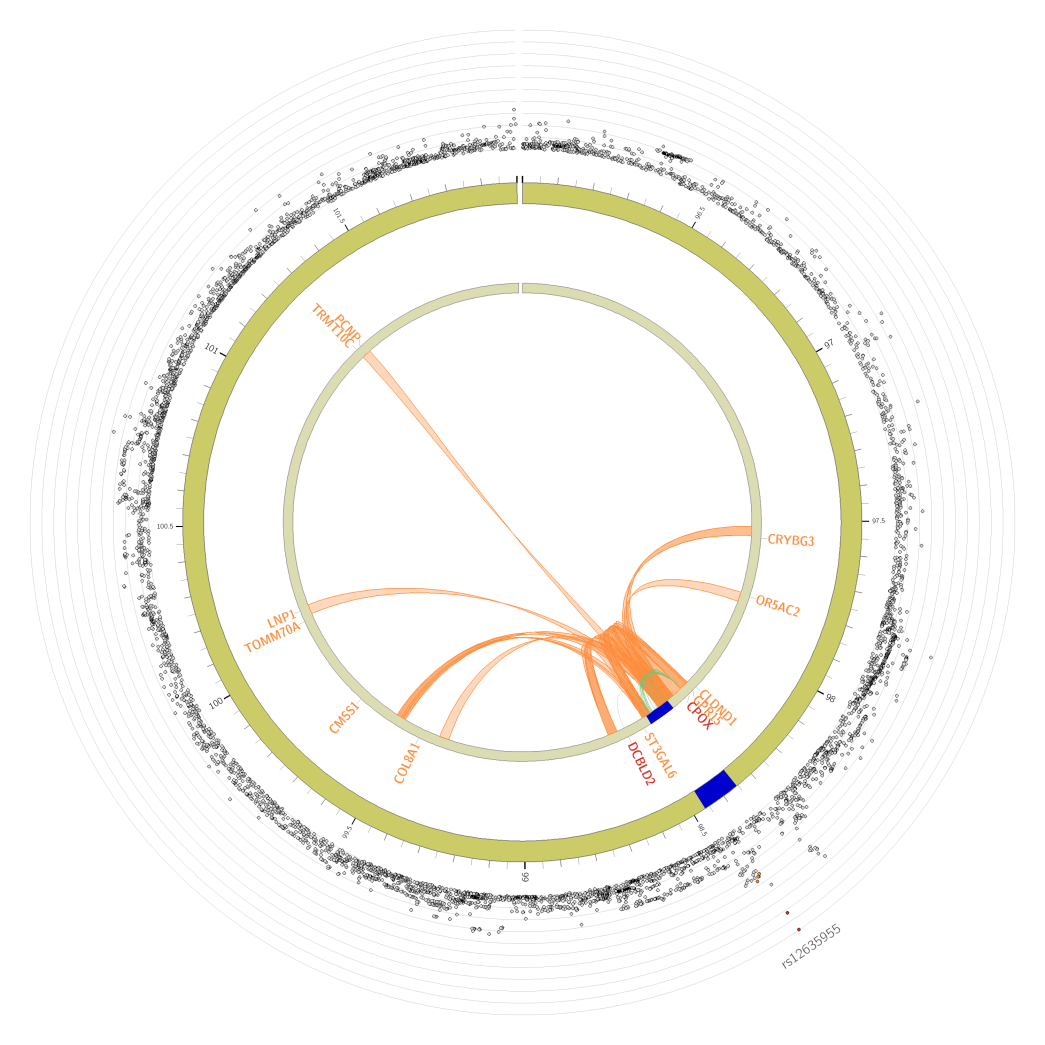 | 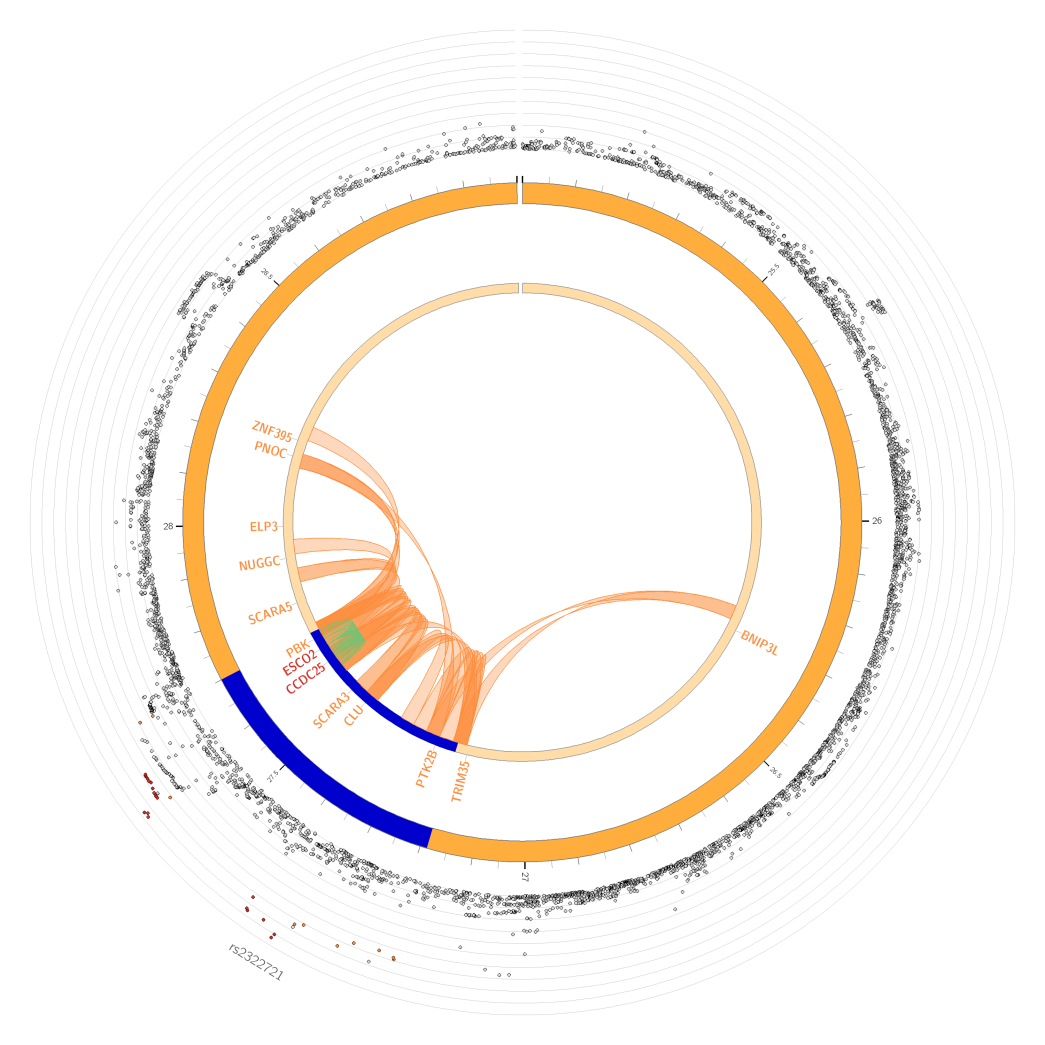 |
| C) Chromosome 3 | D) Chromosome 8 |
| 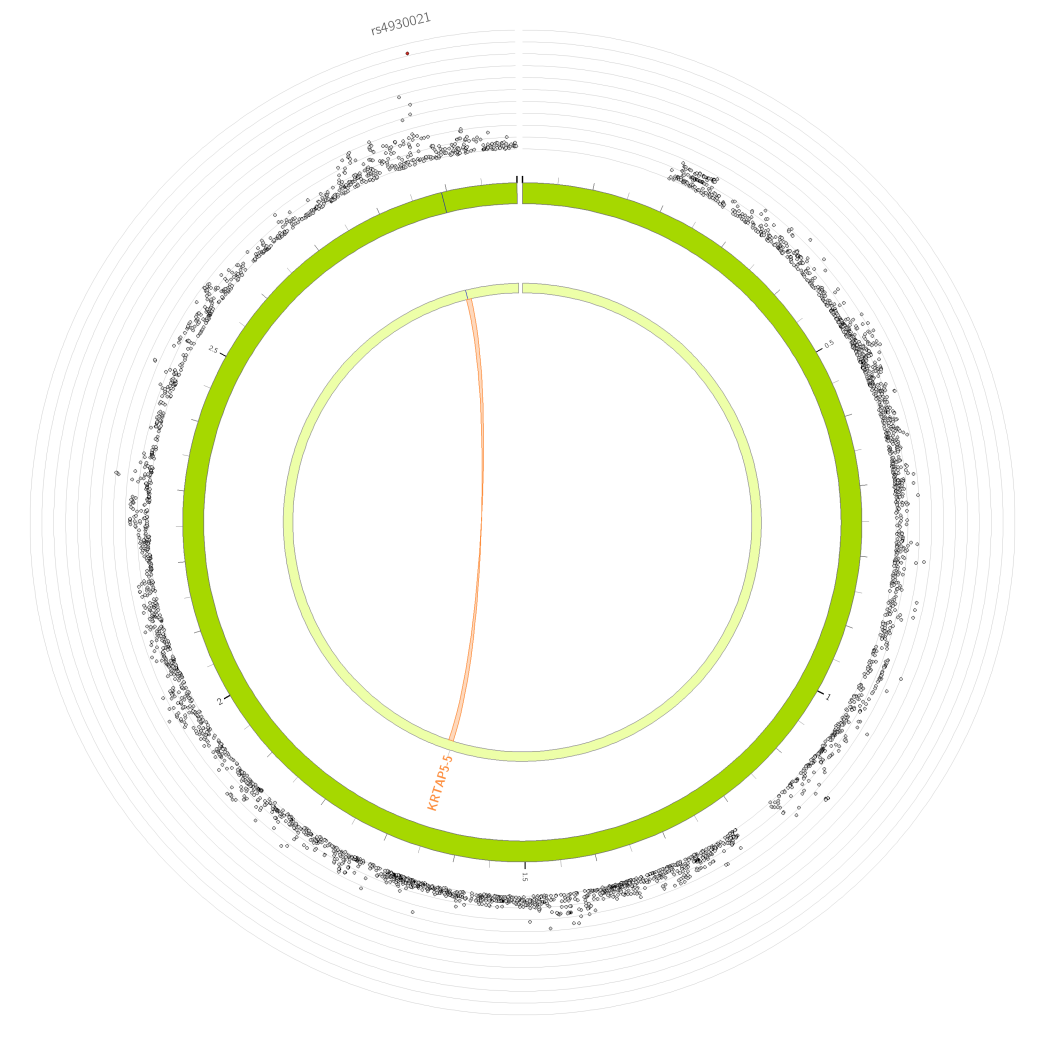 | 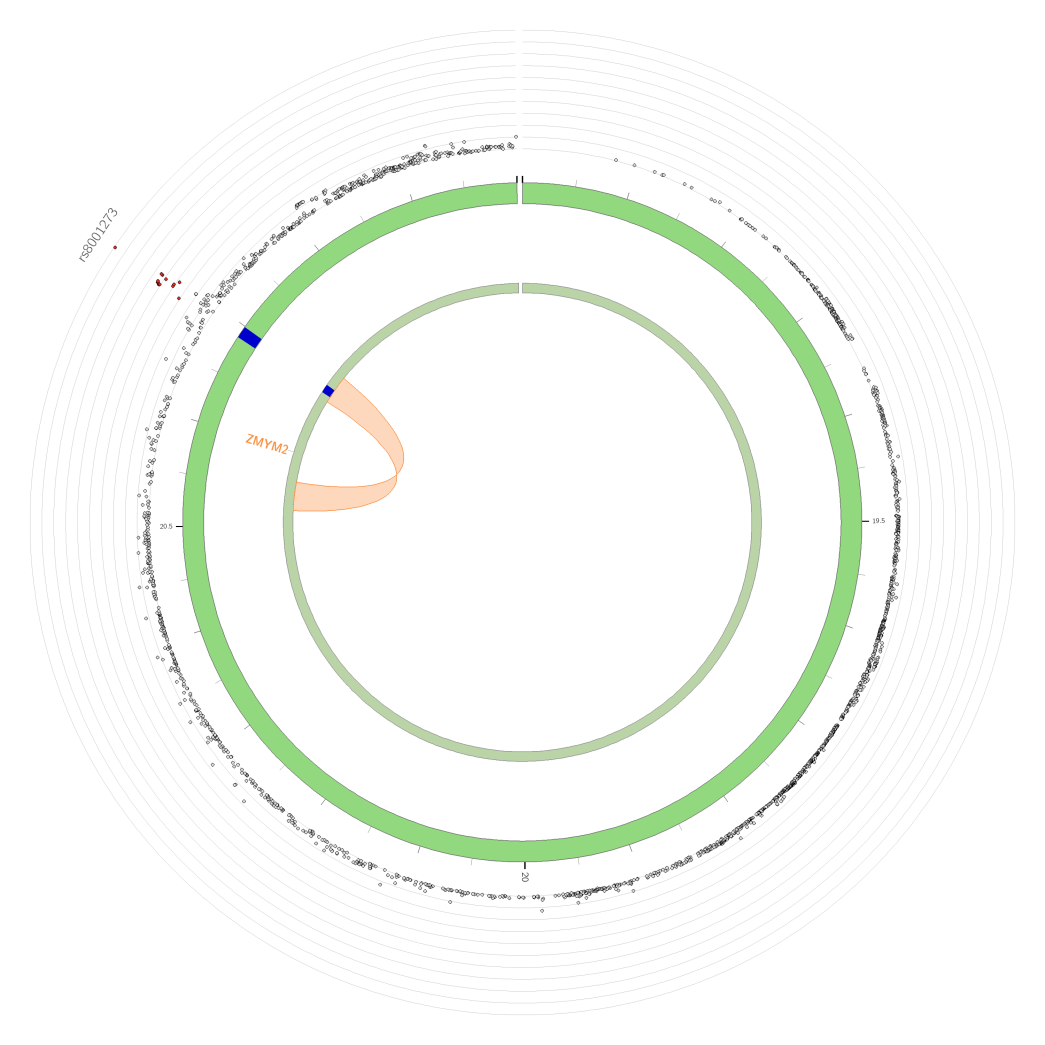 |
| E) Chromosome 11 | F) Chromosome 13 |
| 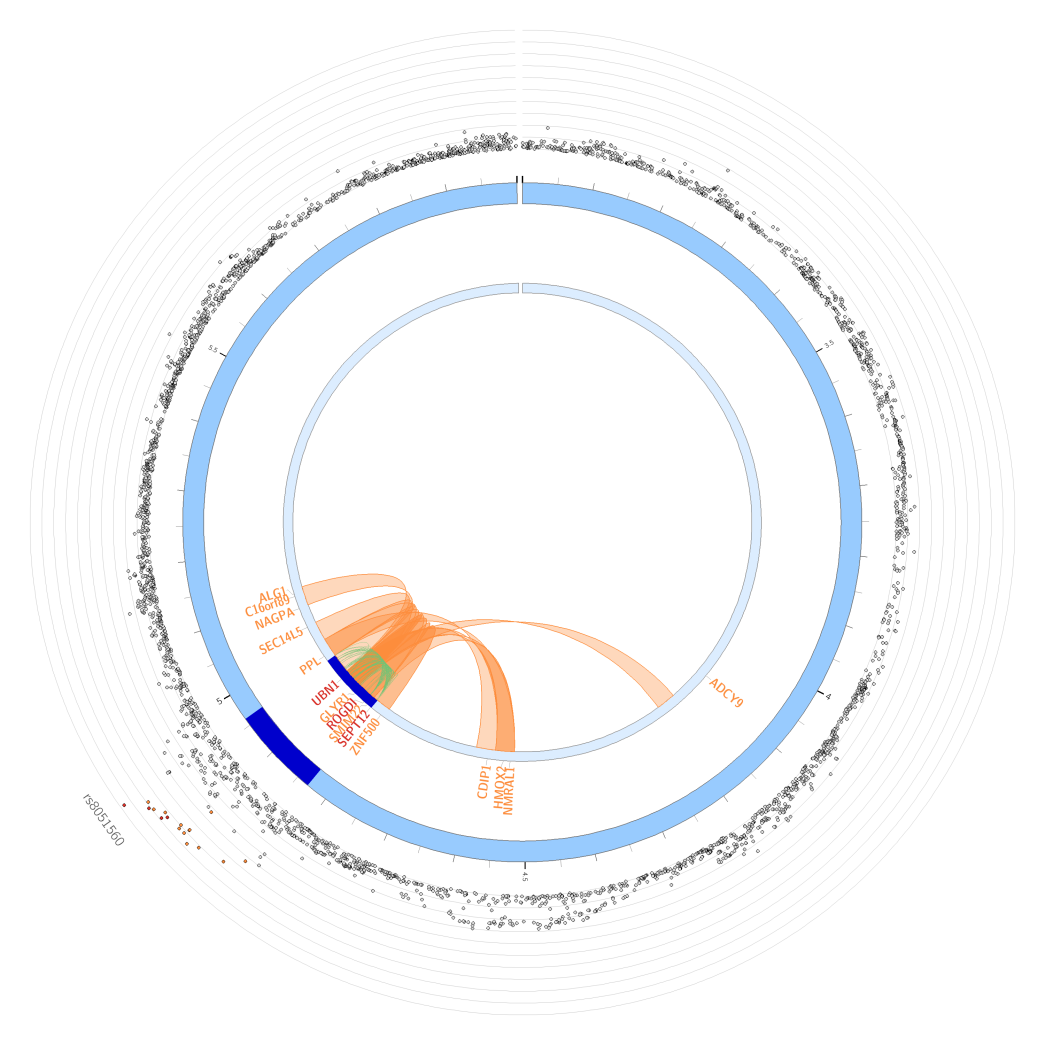 | **Supplementary Figure 5. Circos plot of the chromotin interactions based on stage 3 GWAS result.** A-G were the chromotin interactions in chromosome 1,2,3,8,11,13,16 respectively.   - **Manhattan plot:** The most outer layer. Only SNPs with P < 0.05 are displayed. SNPs in genomic risk loci are color-coded as a function of their maximum r2 to the one of the independent significant SNPs in the locus, as follows: red (r2 > 0.8), orange (r2 > 0.6), green (r2 > 0.4) and blue (r2 > 0.2). SNPs that are not in LD with any of the independent significat SNPs (with r2 ≤ 0.2) are grey.The rsID of the top SNPs in each risk locus are displayed in the most outer layer. Y-axis are raned between 0 to the maximum -log10(P-value) of the SNPs. - **Chromosome ring:** The second layer. Genomic risk loci are highlighted in blue. - **Mapped genes by chromatin interactions or eQTLs:** Only mapped genes by either chroamtin interaction and/or eQTLs are displayed. If the gene is mapped only by chromatin interactions or only by eQTLs, it is colored orange or green, respectively. When the gene is mapped by both, it is colored red. - **Chromosome ring:** The third layer. This is the same as second layer but without coordinates to make it easy to align position of genes with genomic coordinate. - **Chromatin interaction links:** Links colored orange are chromatin interactions. - **eQTL lilnks:** Links colored green are eQTLs. |
| G) Chromosome 16 |  |

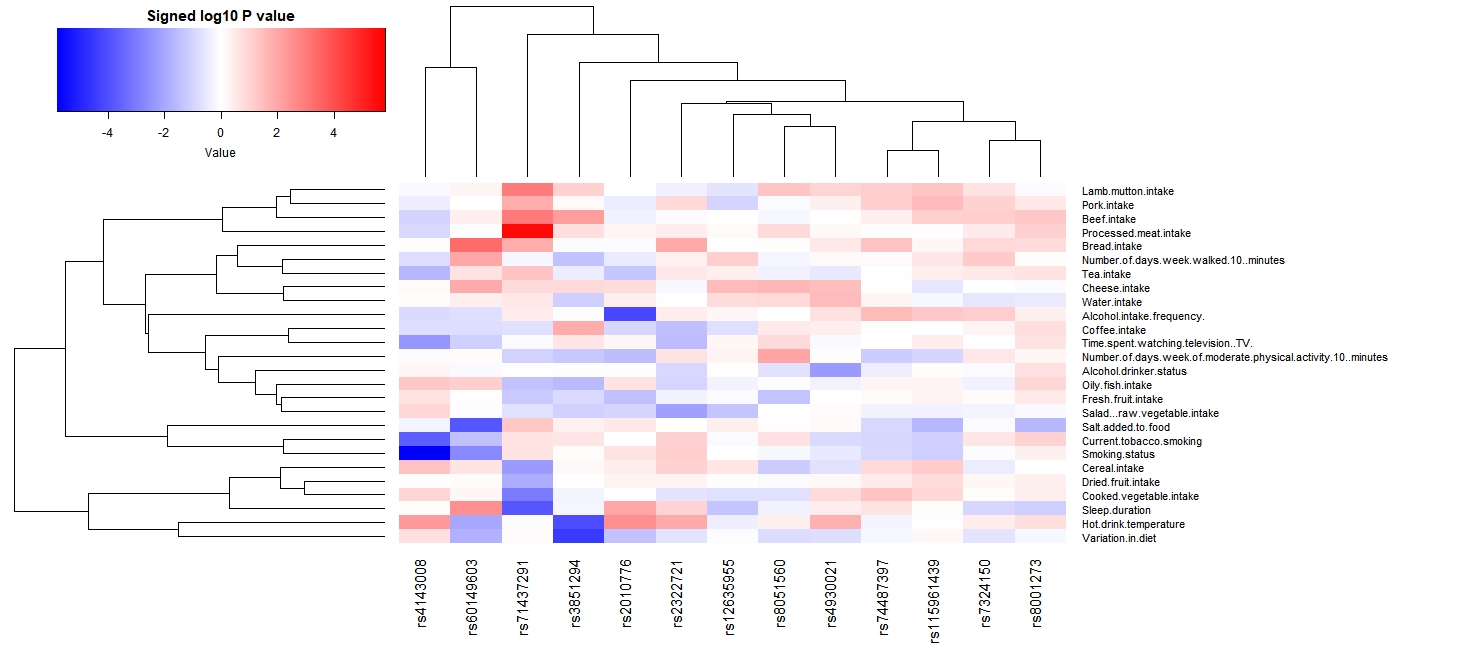

**Supplementary Figure 6.** **Heatmap of RAG loci based on associations with lifestyle-related factors.** For the sentinel SNP at each RAG locus (x-axis), we calculated the -log10(P)*sign(β) (aligned to RAG raising allele) as retrieved from the Gene Atlas catalogue (http://geneatlas.roslin.ed.ac.uk). Red squares indicate direct associations with the trait of interest and blue squares inverse associations. Only SNPs with at least one association at P < 0.05 with at least one of the traits examined are annotated in the heat-map.

| 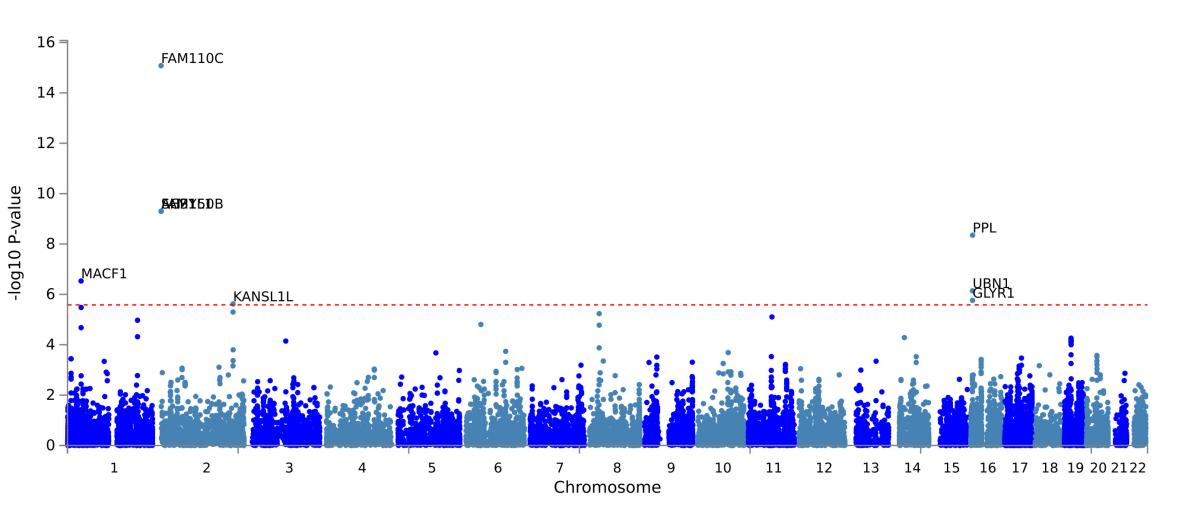 | A |
| --- | --- |
| 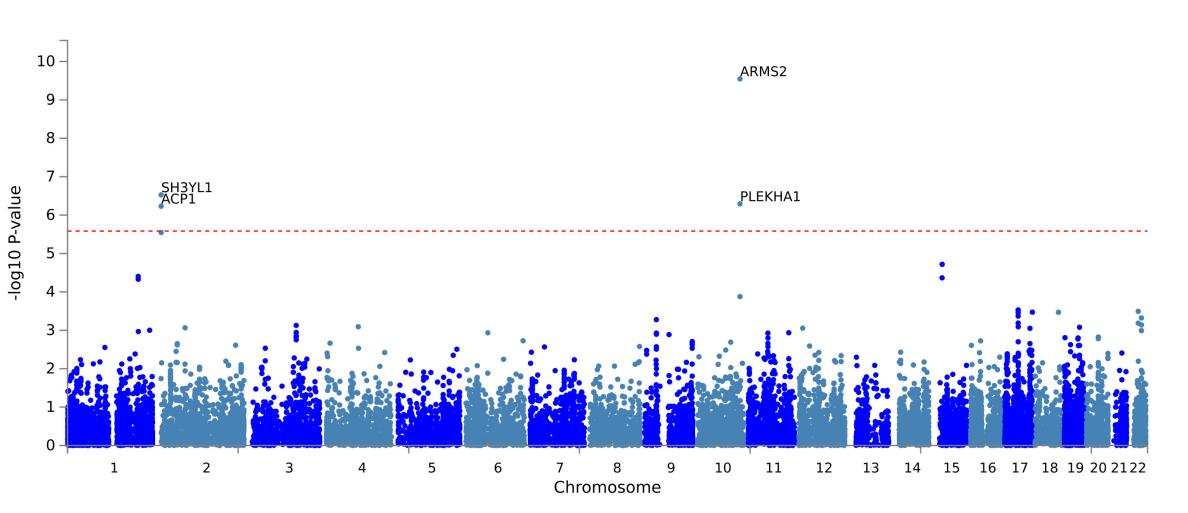 | B |
| 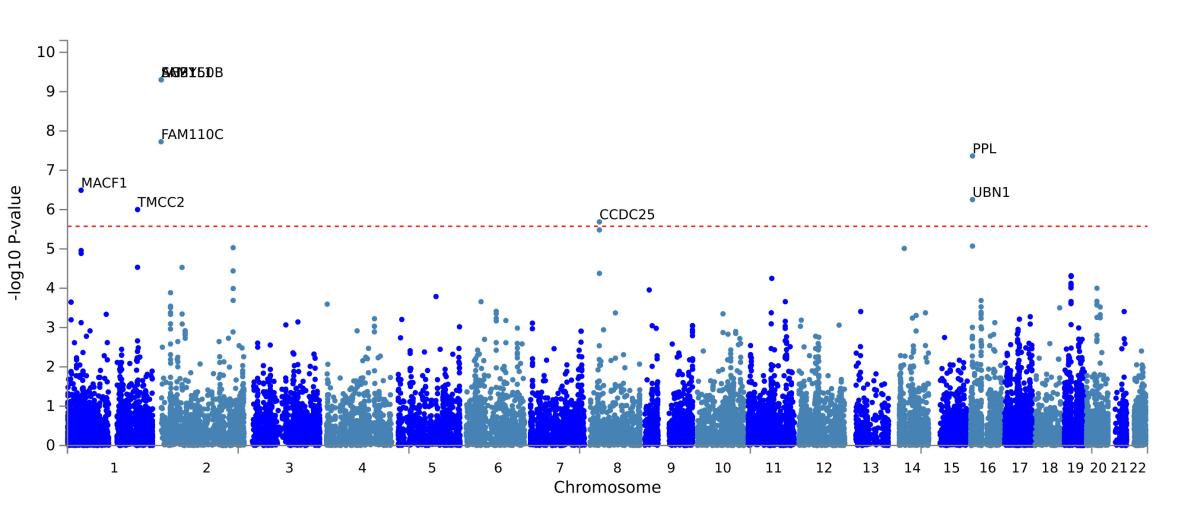 | C |

**Supplementary Figure 7. Gene-based enrichment analysis results.** Gene-based enrichment analysis performed by MAGMA for : A) Stage 1 GWAS results; B) stage 2 GWAS results & C) stage 3 GWAS results. Each dot represents a gene, the x-axis shows the chromosomes where each gene is located, and the y-axis shows −log10 P-value of the enriched gene. The dash horizontal line shows the genome-wide significant threshold (P-value = 2.607e-6 [0.05/19182] ).

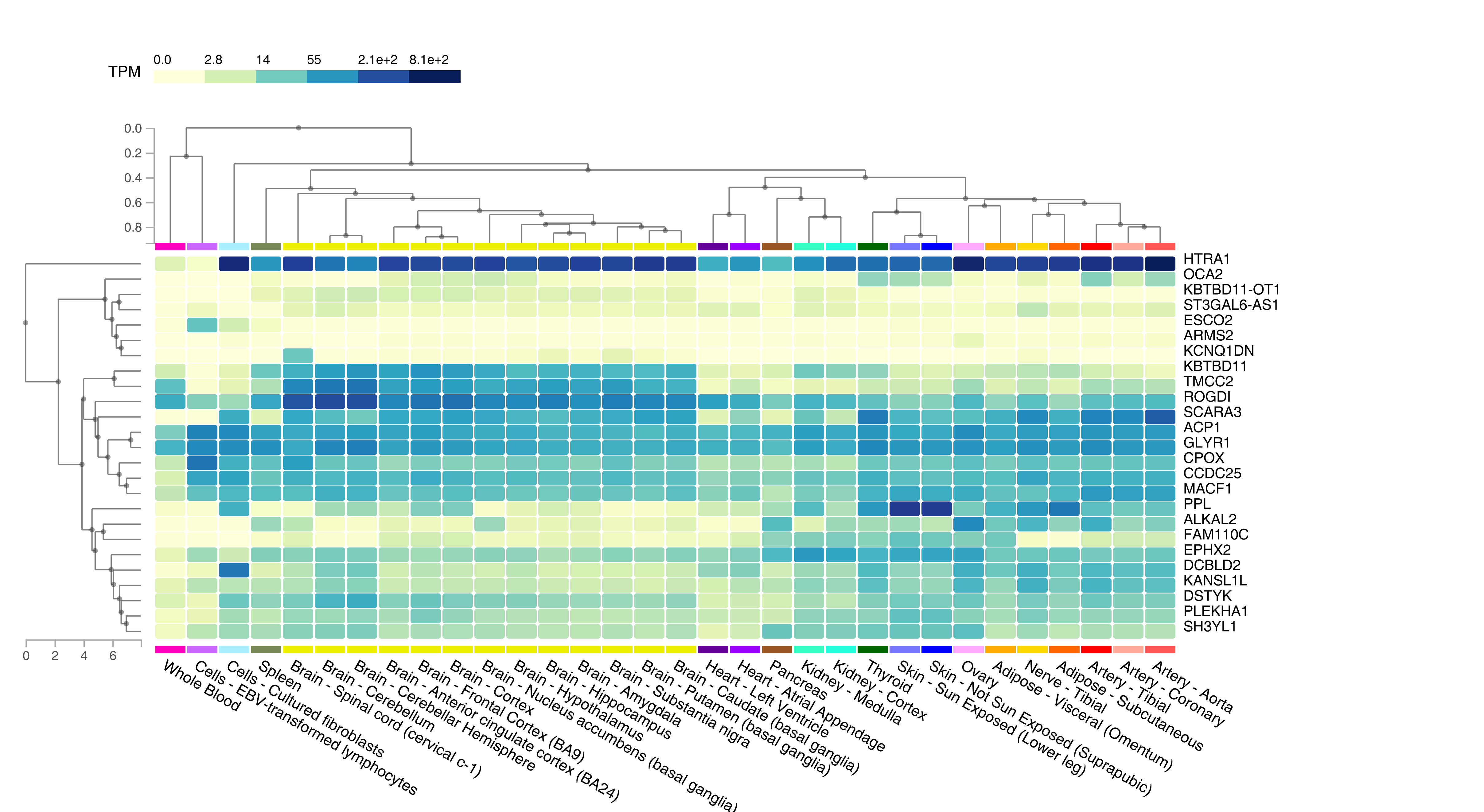
**Supplementary Figure 8. Gene expression level in different tissue.**

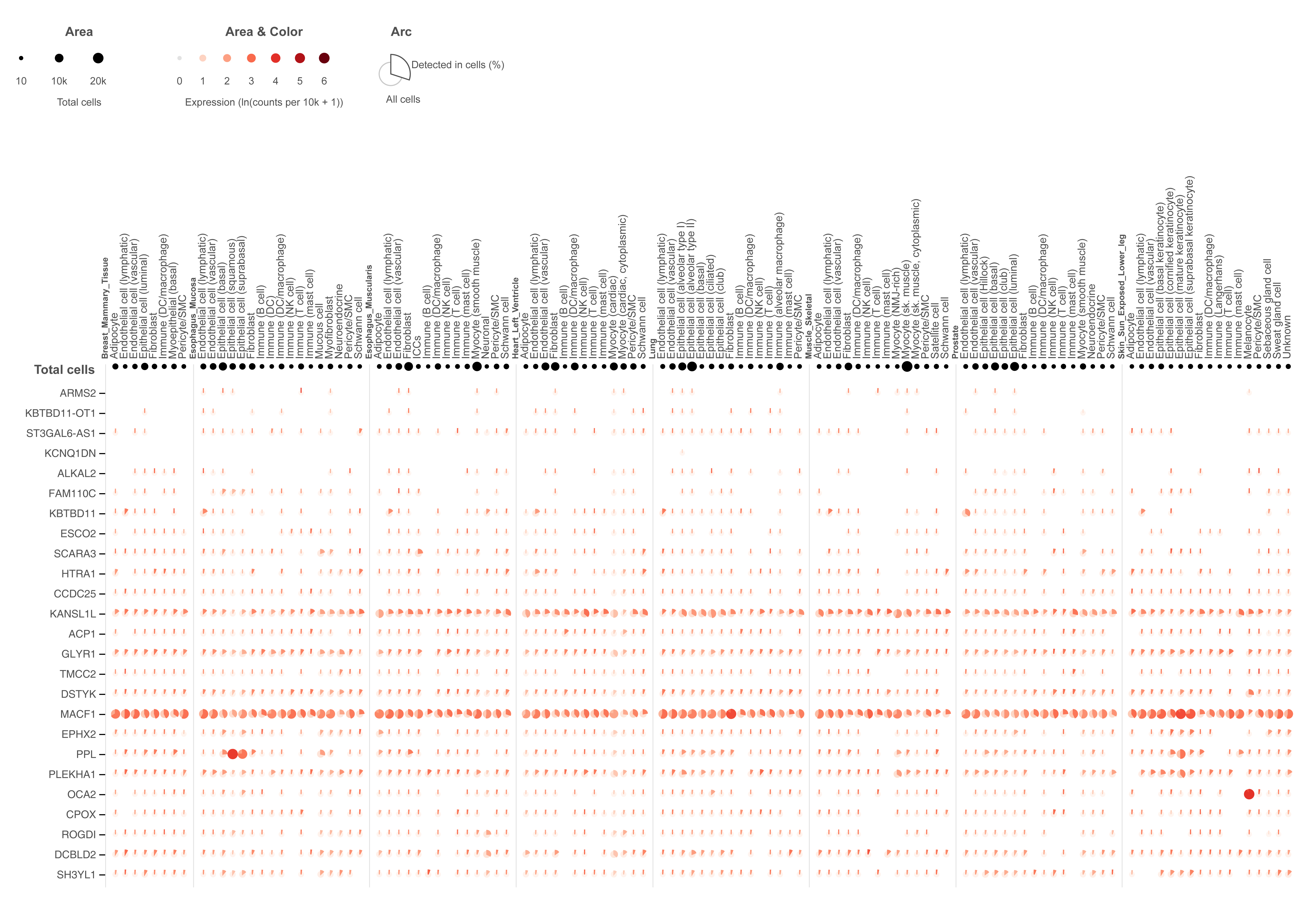

**Supplementary Figure 9. Gene expression level in single cells.**
