## Supplementary Information for "Genomic Determinants of Biological Age Estimated By Deep Learning Applied to Retinal Images"

**Supplementary information -** **Supplementary methods:**

**Supplementary method 1: Ascertain of retinal age within GoDARTS**

Retinal images were pre-processed by the following steps: 1), truncating the excess black, non-retinal regions of the images; 2) re-sizing the image to 260x260 pixels^1^ ; 3), equalizing images on each of the R, G, and B color channels separately; 4), normalizing the pixel intensities were normalized to [0, 1] scale.

Then, EfficientNet-B2^1^ network was used to predict the retinal age. A global average pooling layer, followed by a single output node with linear activation, was used to replace the fully connected layer. All the convolutional layers were pre-trained on ImageNet and were kept for further use.

A 5-fold cross validation strategy was utilized to train the network. The 8,570 individuals with 102,082 images were randomly sampled into 5 folds with 714 individuals (20%) in each fold. Retinal images from people in 4 of the 5 folds were used as 70% training data and 10% validation data in each iteration. The remaining 20% of people in the remaining fold were used for testing. Finally, retinal age prediction was applied for all test photos, totaling 102,082 images.

**Supplementary method 2: Genotyping and imputation process of the UK Biobank cohort**

For the UK biobank, genotype data were available for 488,377 participants. Details of the genotyping and imputation process are in supplementary information. BiLEVE Axiom array, or the UK Biobank Axiom array was used for genotyping by Affymetrix. Prior to data release, UK Biobank researchers performed genotype imputation using a Haplotype Reference Consortium, followed by extensive quality control^2^.

GoDARTS, 25,409 participants underwent genotyping process which were conducted on the Affymetrix 6.0, Illumina Human Omni Express or Broad array in three separated phases. The raw genotype data was imputed separately on the basis of Haplotype Reference Consortium (HRC) reference panel details of the^3^.

**Supplementary method 3: Defining RAG as binary trait**

In our study, as the mean absolute error (MAE) of predicted retinal age was 3.5 years^4^ , we consider our predicted retinal age was comparative more accurate than age predicted basing on DNA methylation clock (MAE = 3.3–5.2 years)^5,6^, blood profile (MAE = 5.5–5.9 years) ^7,8^ or neuroimaging (MAE = 4.3 - 7.3 years)^9^. To select participants with extreme phenotype, we choose people whose RAG is greater or smaller than the MAE. Hence, in our study, participants with a RAG > 3.5 were considered as cases and participants with RAG < 3.5 were consider as controls.

**Supplementary method 4: Identifying patients that are diagnosed as T2D in the UK Biobank**

To identify participants who were diagnosed as type 2 diabetes (T2D), hospital admission information at the baseline was used. Subjects were selected by their ICD10 and ICD 9 (international classification of diseases) code and self-reported information. From the UK Biobank data set, data field ‘41270’, ‘40001’ and ‘40002’ were selected to define participants as T2D by ICD-10 code ‘E10-E14’. Data field ‘41270’, ‘41203’ and ‘41271’ were also selected to define participants who were diagnosed as T2D by ICD9 code defined as '250'. Finally, self-reported medical history collected from touchscreen question "Has a doctor ever told you that you have diabetes?" was also used to define diabetic history and data field ‘2443’ was used. Altogether, 3279 participants with T2D as well as with RAG information were included for the sensitivity GWAS analysis.

**Supplementary information - Supplementary results:**

**Supplementary results 1: Functional annotation of the sentinel 13 SNPs identified by MTAG**

To explore the biological relevance of the identified risk loci, their roles in regulation of gene expression, as well as chromatin interactions were examined. Approximately 79% (154 out of 194) of the sentinel SNPs or those in high LD (r2 > 0.8) have also been reported to be significant expression quantitative trait loci (eQTLs) (FDR < 0.05) in retinal tissue, and these eQTLs were mainly affecting the expression of genes in chromosome 1 (*DSTYK*), chromosome 2 (*SH3YL1*), chromosome 3 (*DCBLD2, CPOX*), chromosome 8 (*CCDC25, ESCO2*) and 16 (*ROGDI, UBN1, SEPT12*) (Supplementary table 2,).

As chromatin interactions information in retinal tissues was not available, only information from neuron tissue (hippocampus, spleen, fetal_cortex, adult cortex, dorsolateral prefrontal cortex), vascular tissue (left ventricle, right ventricle, aorta) embryonic tissues (mesendoderm) and stem cells (mesenchymal stem cell, trophoblast-like cell, hESC, neural progenitor cell) were included for analysis. In different tissues and cell lines, 2016 significant (FDR < 1e-6) chromatin interactions with the leading regions were identified, chromatin interactions were observed in most of the chromosomes (Supplementary table 3, supplementary figure 5).

**Supplementary results 2: RAG SNPs are associated with life style exposures**

By looking up the PheWAS in UK Biobank via Gene ATLAS data base (http://geneatlas.roslin.ed.ac.uk), we explore the association of 13 sentinel SNPs and their associations with lifestyle traits. At a moderate level (P < 0.05), we found genetic associations of RAG variants with different lifestyle traits. Especially for SNP rs71437291 which associated with less dried fruit, cereal, cooked vegetable intake and less sleep duration but more bread, processed meat, red meat intake (all P < 0.015). In general, greater processed meat intake and bread intake are more likely to be associated with increased RAG while some traits such as ‘hot drink temperature’ had heterogeneous effect with RAG. We note that SNP rs4143008 associated with increased RAG was associated with reducing of smoking, which is in counter to the epidemiological expectation (P = 1.64E-06). All these findings might suggest heterogeneous effect of life style on RAG (Supplementary table 5, supplementary figure 6).
